# Clustering-based COPD Subtypes Have Distinct Longitudinal Outcomes and Multi-omics Biomarkers

**DOI:** 10.1101/2022.01.11.22268818

**Authors:** Andrew Gregory, Zhonghui Xu, Katherine Pratte, Sool Lee, Congjian Liu, Robert Chase, Jeong H. Yun, Aabida Saferali, Craig P. Hersh, Russell P. Bowler, Edwin K. Silverman, Peter J. Castaldi, Adel Boueiz

## Abstract

**Introduction:** Chronic obstructive pulmonary disease (COPD) can progress across several domains, complicating the identification of the determinants of disease progression. In our previous work, we applied k-means clustering to spirometric and chest radiologic measures to identify four COPD-related subtypes: “Relatively resistant smokers (RRS)”, “mild upper lobe predominant emphysema (ULE)”, “airway-predominant disease (AD)”, and “severe emphysema (SE)”. In the current study, we examined longitudinal spirometric and radiologic emphysema changes and prospective risks of COPD exacerbations, incident comorbidities, and mortality of these clusters. We also compared their associations to protein and transcriptomic biomarkers.

**Methods:** We included 8,266 non-Hispanic white and African-American smokers from the COPDGene study. We used linear regression to investigate associations to five-year prospective changes in spirometric and radiologic measures and to plasma protein and blood gene expression levels. We used Cox-proportional hazard modeling to test for associations to prospective exacerbations, comorbidities, and mortality.

**Results:** The RRS, ULE, AD, and SE clusters represented 39%, 15%, 26%, and 20% of the studied cohort at baseline, respectively. The SE cluster had the greatest 5-year FEV_1_ and emphysema progression, and the highest risks of exacerbations, cardiovascular disease (CVD), and mortality. The AD cluster had the highest diabetes risk. After adjustments, only the ULE and AD clusters had elevated CVD mortality risks, while only the ULE cluster had the highest cancer-related mortality risk. These clusters also demonstrated differential protein and gene expression biomarker associations.

**Conclusion:** COPD k-means subtypes demonstrate varying rates of disease progression, prospective comorbidities, mortality, and associations to proteomic and transcriptomic biomarkers.

**Funding Sources:** This work was supported by NHLBI K08 HL141601, K08 HL146972, R01 HL116931, R01 HL124233, R01 HL126596, R01 HL116473, U01 HL089897, R01 HL147326, R01 HL130512, and U01 HL089856. The COPDGene study (NCT00608764) is also supported by the COPD Foundation through contributions made to an Industry Advisory Committee comprised of AstraZeneca, Bayer, Boehringer-Ingelheim, Genentech, GlaxoSmithKline, Novartis, Pfizer, Siemens, and Sunovion.

## Introduction

Chronic obstructive pulmonary disease (COPD) is a heterogeneous disorder, with a wide variety of clinical manifestations ^1^. Additionally, disease progression in COPD occurs across multiple domains, such as lung function decline, worsening of emphysema, and development of comorbidities ^2-4^. These challenges complicate COPD subtyping and the identification of the determinants of COPD progression.

Several reports have shown that factors, such as COPD exacerbation history, reduced pulmonary function, and a low BMI, are associated with an elevated risk of respiratory exacerbations, accelerated spirometric decline, and emphysema changes ^2 5 6^. Additionally, studies which have subtyped subjects with COPD based on spirometry, respiratory symptoms and other characteristics have revealed that these subgroups differ in their risks for exacerbations, hospital admissions, and FEV_1_ and emphysema changes ^7-11^. However, many of these studies have modest sample size and limited longitudinal follow-up. Furthermore, while comorbid conditions such as cardiovascular disease (CVD) and type 2 diabetes mellitus are common in subjects with COPD, the specific COPD-related characteristics associated with the risk of developing these comorbidities have not been fully described ^12 13^. Moreover, different protein biomarkers have been identified in subjects with COPD, but few studies have assessed subtype-specific proteomic signatures ^14^. Similarly, while transcriptomic signatures have been identified in COPD, the differences in such patterns between COPD subtypes have not been thoroughly investigated ^15-17^.

In our previous work, we applied k-means clustering to spirometric and chest radiologic measures and identified four COPD-related subtypes: “Relatively resistant smokers (i.e. no/mild airflow obstruction and minimal emphysema despite heavy smoking) (RRS)”, “mild upper lobe predominant emphysema (ULE)”, “airway-predominant disease (AD)”, and “severe emphysema (SE)”, which had differing cross-sectional profiles and genetic associations ^18^. In the current study, we included up to 12.7 years of prospective data to investigate subtype-specific rates of progression in spirometric measures and radiologic emphysema and to quantify the risks of prospective COPD exacerbations, CVD events, diabetes, and mortality. We also investigated cross-sectional associations between subtypes and plasma protein and blood transcriptomic biomarkers. We hypothesized that these subtypes would have different disease progression profiles and associations to biomarkers. Some of these results have been previously reported as an abstract ^19^.

## Methods

### Study description

The COPDGene study is a prospective, multicenter, longitudinal study investigating the genetic and epidemiological characteristics of COPD across 21 centers in the U.S. (NCT00608764, www.copdgene.org) ^20^. Institutional review board approval was obtained at each study center. Patients or the public were not involved in the design, conduct, reporting, or dissemination plans of our research. All data produced in the present study are available upon reasonable request to the authors. This study enrolled non-Hispanic whites and African-Americans, who were 45-80 years old and had at least 10 pack-years of lifetime smoking history. Subjects were recruited across the full spectrum of disease severity as defined by the Global Initiative for Chronic Obstructive Lung Disease (GOLD) spirometric grading system ^20^. COPDGene conducted two study visits (Visit 1 and Visit 2) approximately 5 years apart. Subjects were also contacted every 3-6 months through the COPDGene Longitudinal Follow-up program via phone or online surveys to collect data on incident COPD-related events, comorbidities, and mortality. COPD-related events and comorbidities included self-reported COPD exacerbations (defined as the acute worsening of respiratory symptoms that required systemic steroids and/or antibiotics ^21^), CVD events (defined as a composite endpoint of stroke, heart attack, coronary artery disease, coronary artery bypass graft surgery, peripheral artery disease, and/or cardiac angina), and type 2 diabetes mellitus. All-cause mortality was determined through a combination of longitudinal follow-up and a search of the social security death index. Cause-specific mortality was categorized as respiratory-related, CVD-related, cancer-related (any type) or due to other causes and determined through systematic adjudication process based on the methods used in the Towards a Revolution in COPD Health (TORCH) trial ^22^.

Demographics, spirometry, imaging, smoking burden, respiratory symptoms, and comorbidities were collected at Visits 1 and 2. In addition to GOLD grades 0-4, we included subjects with Preserved Ratio Impaired Spirometry (PRISm), defined as FEV_1_/FVC ≥ 0.70 but with FEV_1_ < 80% predicted ^21^. Thirona software (www.thirona.eu) was used to quantify emphysema as the percentage of lung voxels with an attenuation of < -950 HU at maximal inspiration (%LAA-950) ^23^. The Hounsfield units at the 15^th^ percentile of the computed tomography (CT) density histogram at end-inspiration corrected for the depth of inspiratory variation (adjusted Perc15 density) were used for longitudinal changes in emphysema ^24 25^. Per convention, adjusted Perc15 density values are reported as HU + 1000. The levels of 1,305 protein biomarkers (SOMAscan Human Plasma 1.3K assay) were obtained from plasma samples collected at Visit 1 ^26^. Total blood RNA was collected at Visit 2.

### Cluster Generation

We used the k-means clusters that were generated in our previously published work ^18^. FEV_1_ percent predicted, CT-quantified emphysema, percent airway wall thickness, and apico-basal emphysema distribution (log of the lung upper third to lower third ratio of emphysema) were the input features that were used for clustering at Visit 1. Using the same approach, we also performed k-means clustering at Visit 2 to assess cluster assignment stability between the two visits and to conduct differential gene expression analyses using RNA-Seq data available at Visit 2.

### Statistical analyses

Data distributions were reported as medians with interquartile ranges or counts with percentages, where appropriate. We calculated FEV_1_ and emphysema changes as either absolute or relative annualized changes. We computed absolute annualized changes by subtracting Visit 1 from Visit 2 values and dividing the difference by the time in years between visits for each subject. Relative annualized changes were obtained by dividing the absolute annualized changes by Visit 1 values and multiplying by 100. Negative values indicate worsening of the disease between visits. We used the Kruskal-Wallis and chi-square tests for continuous and categorical variables, respectively. We subsequently performed post-hoc pairwise comparisons between the clusters using the Nemenyi and chi-square tests for continuous and categorical variables, respectively. Additionally, we constructed univariable and multivariable linear regression models to relate changes in FEV_1_ and emphysema as well as plasma proteins to cluster assignment. We assessed risks of incident COPD exacerbations, CVD events, diabetes, and mortality using Cox proportional hazards models and obtained survival curves using the Kaplan-Meier method. For the analyses of incident CVD events and diabetes, we excluded subjects who had a history of CVD or diabetes at Visit 1. We used the RRS cluster as the reference group. Linear regression and Cox models were adjusted for relevant baseline physiological, clinical, and demographic characteristics. We additionally adjusted for metabolic syndrome in the CVD multivariable Cox models and for body mass index, airflow obstruction and exercise capacity (BODE index) in the mortality models. For the protein analyses, we adjusted for age, sex, race and current smoking status. To ensure that cluster associations to emphysema changes are not confounded by CT scanner type, a sensitivity analysis was performed by adding scanner type as a covariate in a subgroup analysis limited to subjects who underwent scans with the same scanner type between visits.

To test for differential gene expression between clusters, we used the linear modeling approach implemented in the limma R package (v3.38.3) ^27^ adjusting for age, sex, current smoking, white blood cell count proportions, and library prep batch. Gene ontology (GO) functional enrichment of the gene sets was calculated using the weighted Fisher test in the topGO Bioconductor package that accounts for the dependency between terms in the GO topology ^28^. We reported only the GO pathways with at least 3 significant genes. All tests for the clinical outcomes were two-tailed with a significance threshold of P-value < 0.05. For the protein and RNA-Seq analyses, we corrected for multiple comparisons using the Benjamini-Hochberg method and applied a threshold of significance of false discovery rate (FDR) of 10% ^29^. Significantly enriched GO pathways were identified using the weighted Fisher P-value < 0.005.

Additional methods are available in the Supplement.

## Results

The overview of the study is shown in Figure 1 and the study flow diagram is outlined in Figure S1. Subjects who were not included in the analyses of longitudinal changes in FEV_1_ and emphysema, 935 of whom died between the first and second study visit, had a higher proportion of GOLD spirometric grade 4 disease (Table S1).

**Figure 1.**
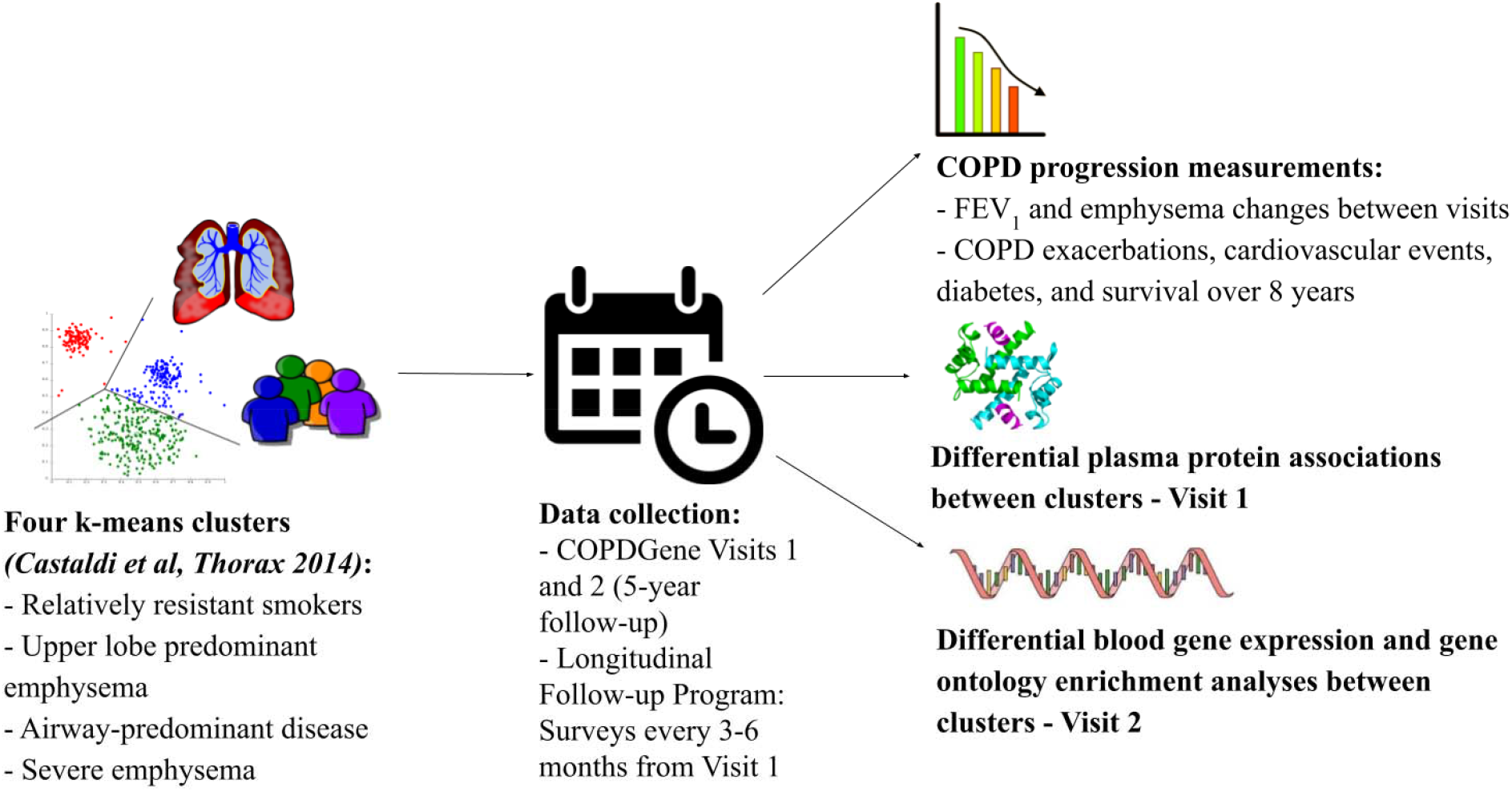
Study design. The goal of the study was to analyze COPD progression, differential plasma protein associations and blood gene expression, and gene ontology (GO) enrichment characteristics of the four clusters that we identified in our previous k-means clustering analysis in the COPDGene study *(Castaldi et al, Thorax 2014)*.

### Cluster characteristics at baseline and stability of cluster assignments between visits

The RRS cluster represented 39% of the studied population at the baseline visit and was characterized by a history of heavy smoking without significant airflow obstruction, emphysema, or airway wall thickness compared to the other clusters *(P-values < 0*.*05)* (Table S2). Additionally, subjects in the RRS cluster had predominantly GOLD 0-1 spirometry. At Visit 2, RRS cluster membership was stable as 76.3% of subjects were still assigned to this cluster (Figure S2). The ULE cluster consisted of 15% of all subjects and had moderate airflow obstruction and mild emphysema predominantly in the upper lung lobes. The ULE cluster was relatively unstable, with only 38% of subjects remaining within this cluster at their follow-up visits. The AD cluster, 26% of all participants, was characterized by high BMI and had the highest proportion of subjects with PRISm *(P-values < 0*.*05)*. The AD cluster was more stable than the ULE cluster with 53% of subjects staying within this cluster at their follow-up visits. The SE cluster, 20% of the studied cohort, exhibited high emphysema and gas trapping and had the highest proportion of GOLD 3-4 smokers *(P-values < 0*.*05)*. The SE cluster was very stable as 92.2% of subjects remained within the SE cluster at Visit 2. For subsequent analyses of longitudinal outcomes and protein data by k-means clusters, subjects were analyzed according to their cluster assignment at Visit 1.

### Cluster-specific rates of spirometric and emphysema progression

The 5-year change values for FEV_1_ (measured as absolute change in FEV_1_ and percent change relative to baseline) and emphysema are shown in Figure 2, and the results from the univariable and multivariable models are shown in Table 1. The RRS cluster, which has the least impaired spirometry and emphysema at baseline, had the greatest loss in absolute FEV_1_. The AD cluster had the lowest absolute loss in FEV_1_, significantly less than the RRS cluster in both univariable and multivariable models. Both the ULE and SE clusters had similar absolute FEV_1_ changes relative to the RRS cluster. However, when adjusted for relevant covariates, the SE cluster had significantly less absolute FEV_1_ decline than the RRS cluster. While absolute FEV_1_ changes were notably higher in the RRS cluster, percent changes in FEV_1_ relative to baseline were most pronounced in the SE cluster. Pairwise comparisons between all clusters showed that both emphysema-related clusters (ULE and SE) had significantly larger relative changes in percent FEV_1_ relative to baseline than both the RRS and AD clusters (*P-values < 0*.*05*, Table S3).

**Figure 2.**
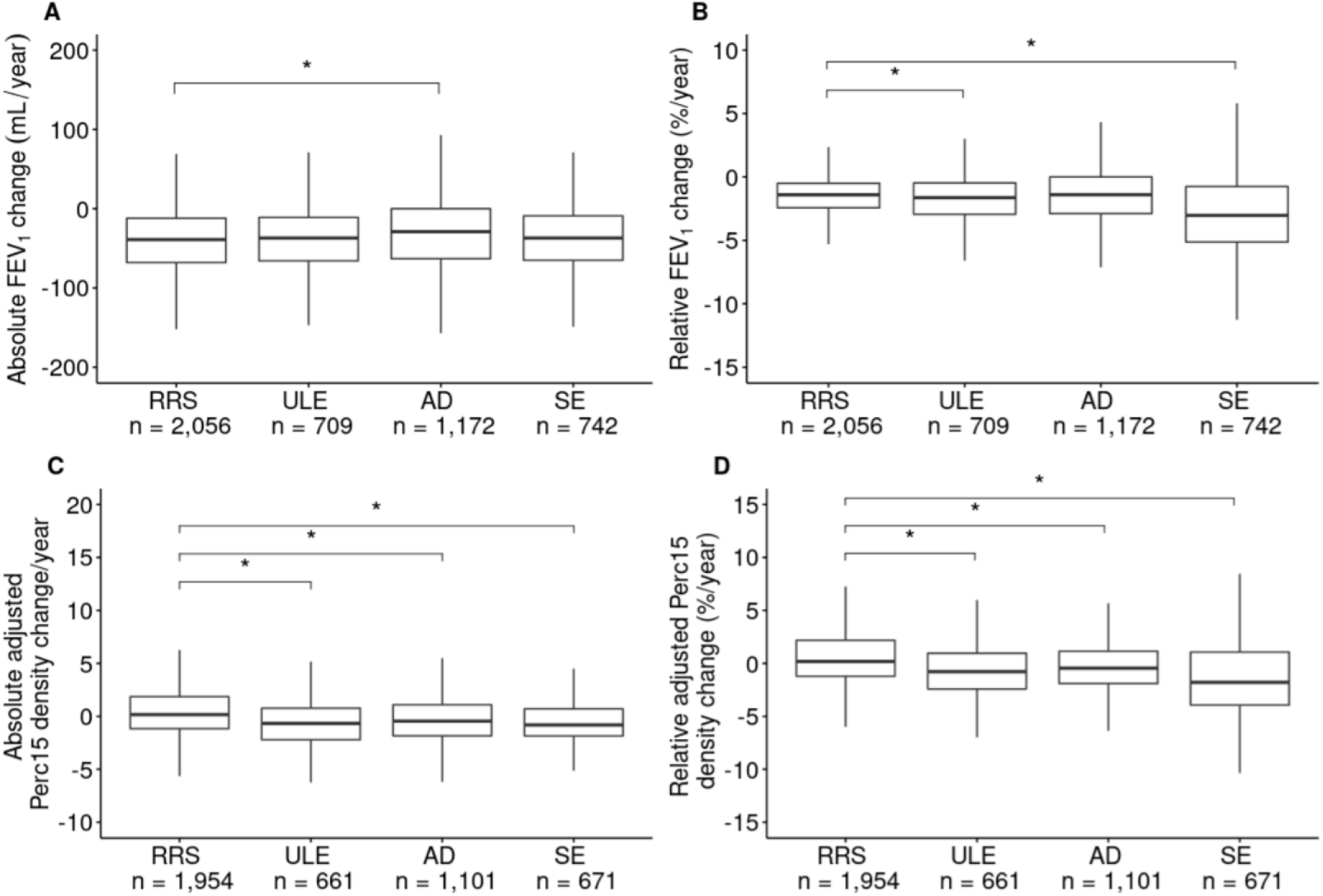
Disease progression by k-means cluster. *(A)* Absolute change in FEV_1_ (mL/year). *(B)* Relative change in FEV_1_ (change as % of baseline value/year). *(C)* Absolute change in emphysema measured as adjusted Perc15 density change/year. *(D)* Relative change in emphysema measured as adjusted Perc15 density change (% of baseline value/year). P-values < 0.05 are indicated by an asterisk. Abbreviations: RRS = Relatively resistant smokers; ULE = Upper lobe predominant emphysema; AD = Airway-predominant disease; SE = Severe emphysema.

**Table 1.**
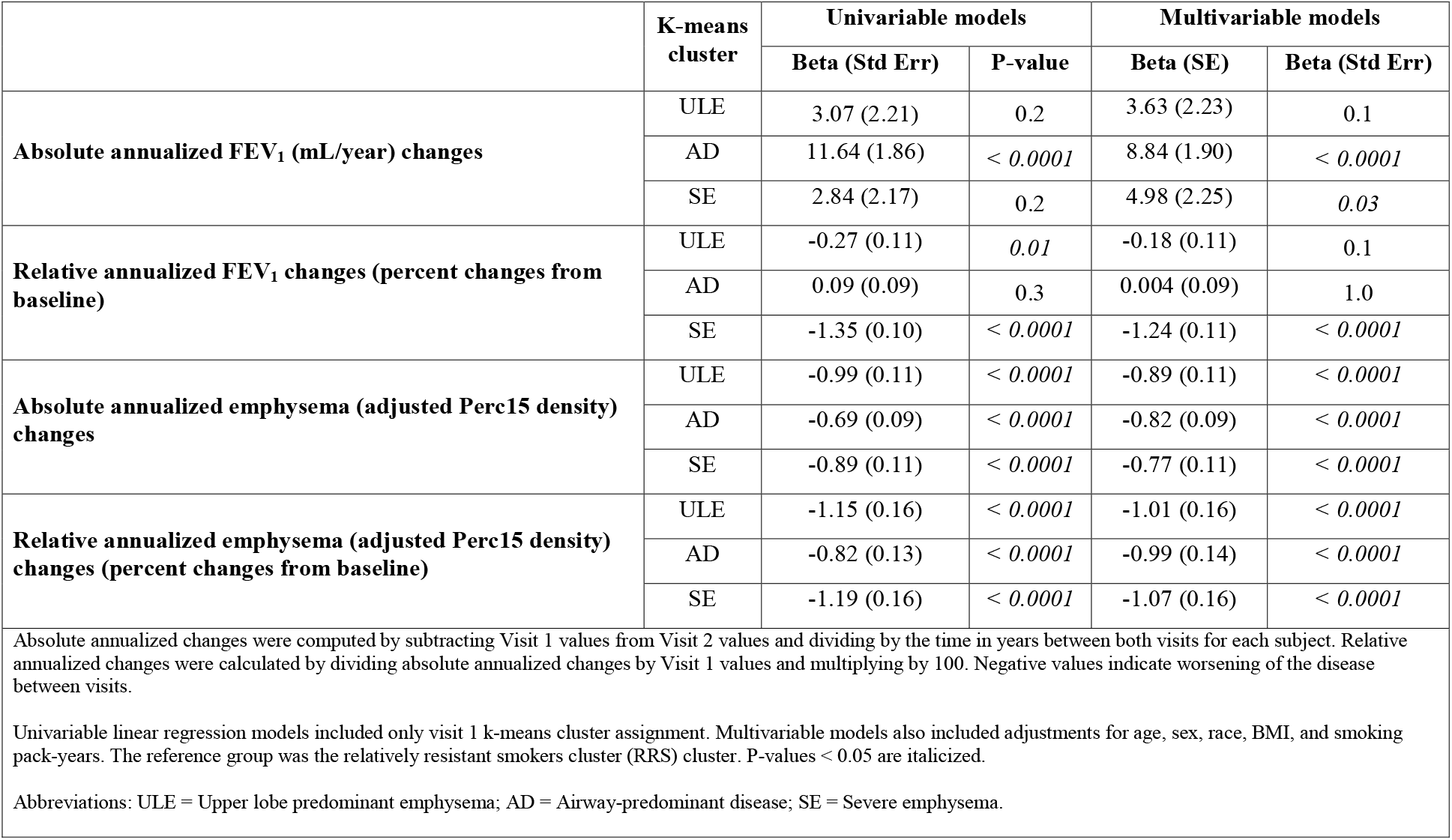
Associations of k-means clusters with absolute and relative annualized FEV_1_ and emphysema changes

|  | K-means cluster | Univariable models |  | Multivariable models |  |
| --- | --- | --- | --- | --- | --- |
|  |  | Beta (Std Err) | P-value | Beta (SE) | Beta (Std Err) |
| **Absolute annualized FEV<sub>1</sub> (mL/year) changes** | ULE | 3.07 (2.21) | 0.2 | 3.63 (2.23) | 0.1 |
|  | AD | 11.64 (1.86) | < 0.0001 | 8.84 (1.90) | < 0.0001 |
|  | SE | 2.84 (2.17) | 0.2 | 4.98 (2.25) | 0.03 |
| **Relative annualized FEV<sub>1</sub> changes (percent changes from baseline)** | ULE | -0.27 (0.11) | 0.01 | -0.18 (0.11) | 0.1 |
|  | AD | 0.09 (0.09) | 0.3 | 0.004 (0.09) | 1.0 |
|  | SE | -1.35 (0.10) | < 0.0001 | -1.24 (0.11) | < 0.0001 |
| **Absolute annualized emphysema (adjusted Perc15 density) changes** | ULE | -0.99 (0.11) | < 0.0001 | -0.89 (0.11) | < 0.0001 |
|  | AD | -0.69 (0.09) | < 0.0001 | -0.82 (0.09) | < 0.0001 |
|  | SE | -0.89 (0.11) | < 0.0001 | -0.77 (0.11) | < 0.0001 |
| **Relative annualized emphysema (adjusted Perc15 density) changes (percent changes from baseline)** | ULE | -1.15 (0.16) | < 0.0001 | -1.01 (0.16) | < 0.0001 |
|  | AD | -0.82 (0.13) | < 0.0001 | -0.99 (0.14) | < 0.0001 |
|  | SE | -1.19 (0.16) | < 0.0001 | -1.07 (0.16) | < 0.0001 |
| Absolute annualized changes were computed by subtracting Visit 1 values from Visit 2 values and dividing by the time in years between both visits for each subject. Relative annualized changes were calculated by dividing absolute annualized changes by Visit 1 values and multiplying by 100. Negative values indicate worsening of the disease between visits.<br>Univariable linear regression models included only visit 1 k-means cluster assignment. Multivariable models also included adjustments for age, sex, race, BMI, and smoking pack-years. The reference group was the relatively resistant smokers cluster (RRS) cluster. P-values < 0.05 are italicized.<br>Abbreviations: ULE = Upper lobe predominant emphysema; AD = Airway-predominant disease; SE = Severe emphysema. |  |  |  |  |  |

Compared to FEV_1_ changes, the pattern of CT-quantified emphysema progression was less sensitive to the metric being used (absolute *vs*. relative). The SE cluster had the most rapid while the RRS cluster had the least rapid relative progression in both univariable and multivariable models. The ULE and AD clusters had significantly greater absolute and relative emphysema changes than the RRS cluster. The ULE cluster had significantly greater relative emphysema progression than the AD cluster (*Pairwise P-value: 0*.*03*, Table S3). We noted similar cluster associations to both absolute and relative emphysema changes when we added scanner type as a covariate in the subgroup analysis limited to subjects who underwent scans with the same scanner type between visits (Table S4).

### Cluster-specific risk of incident co-morbidities and COPD-related events

Starting from Visit 1, the median follow-up time was 9.2 years for prospective CVD and diabetes, 5.3 years for respiratory exacerbations, 9.5 years for all-cause mortality, and and 7.8 years for cause-specific mortality. When we analyzed prospective risks of various health outcomes by subtype, we observed that the SE cluster had the highest risk for prospective COPD exacerbations and incident CVD (Figure 3). After multivariable adjustment, the SE cluster had a 3 times higher likelihood of having a COPD exacerbation when compared to the RRS cluster *(HR 2*.*98 (SE: 0*.*05), P-value < 0*.*001)* (Table S5). The AD cluster had the highest risk of incident diabetes *(HR 1*.*97 (SE: 0*.*09), P-value < 0*.*001)* and this association remained significant after correcting for age, sex, race, BMI, and smoking pack-years. COPD exacerbation and CVD risks were higher in the ULE cluster compared to the RRS cluster (*P-values < 0*.*05*).

**Figure 3.**
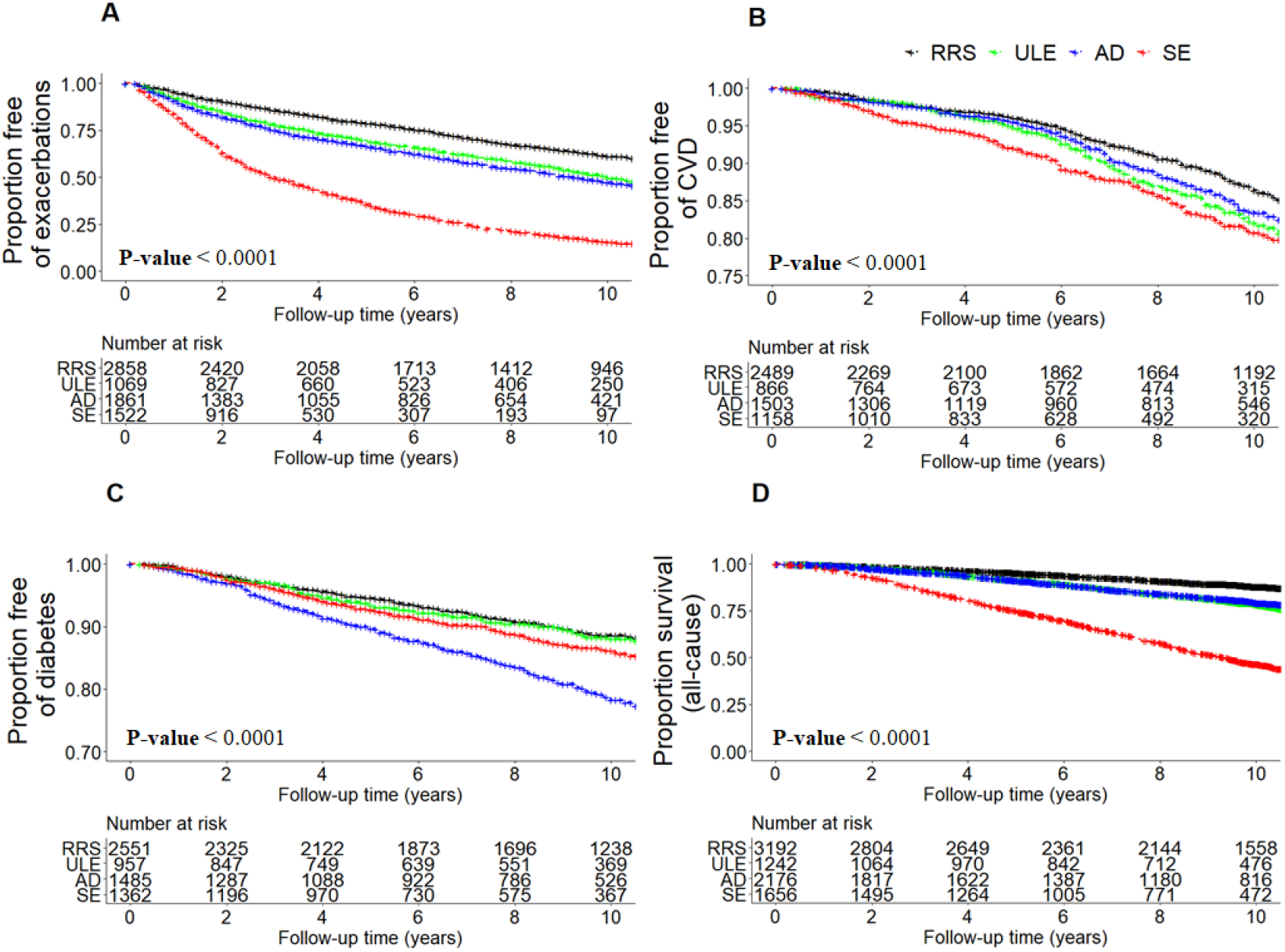
Kaplan-Meier plots of COPD-related events by k-means cluster. *(A)* COPD exacerbation, defined as the acute worsening of respiratory symptoms that required antibiotics and/or systemic steroids. *(B)* Cardiovascular disease (CVD) event, defined as a composite endpoint of stroke, heart attack, coronary artery disease, coronary artery bypass graft surgery, peripheral artery disease, and/or cardiac angina. *(C)* Diabetes. *(D)* All-cause mortality. For CVD events and diabetes, subjects who had a history of CVD or diabetes at Visit 1 were excluded from the analysis. Abbreviations: RRS = Relatively resistant smokers; ULE = Upper lobe predominant emphysema; AD = Airway-predominant disease; SE = Severe emphysema.

Survival curves by subtype are shown for all-cause mortality (Figure 3) and for cause-specific mortality (Figure S3). Results of the survival models are presented in Table S6. In the univariable models, subjects in the SE cluster had the highest risks of all-cause, respiratory-related, CVD-related, cancer-related and other causes-related mortality relative to the RRS cluster (6, 50, 3, 3, and 2 times higher, respectively, *P-values < 0*.*0001*). The risks for all-cause, respiratory, and CVD mortality were also elevated for the ULE and AD clusters relative to RRS *(P-values < 0*.*05)*. In multivariable models adjusting for age, sex, race, smoking pack-years, and BODE, statistical significance was maintained for the associations of the ULE, AD and SE clusters with all-cause mortality, the association of the SE cluster with respiratory mortality, the associations of the ULE and AD clusters with CVD mortality, and the association of the ULE cluster with cancer mortality (*P-values < 0*.*05*).

### Clusters associations to protein and gene expression biomarkers

Using SOMAscan plasma protein measurements at baseline from 1,047 subjects, we tested for differential protein associations between clusters. Using the RRS group as the reference, we identified significant associations in adjusted models to 16, 65, and 219 proteins for the ULE, AD, and SE clusters, respectively *(FDR 10%)* (Table S7). The most strongly associated proteins for the ULE cluster were related to mitochondrial function (ATP synthase peripheral stalk subunit OSCP and glucokinase regulatory protein) and cytoskeleton rearrangement (serine/threonine-protein kinases MRCK beta and PAK 6) (Table 2). Top proteins associations for the AD cluster were primarily involved in fatty acid metabolism, such as elevated fatty acid-binding protein, leptin, and retinoic acid receptor responder protein 2 and decreased apolipoprotein M. For the SE cluster, top associated proteins were related to innate immunity, such as bactericidal permeability-increasing protein, complement component C9, and protein S100-A12. The overlap of the protein associations between subtypes is shown in Figure S4.

**Table 2.** Top 5 unique significantly differentially associated proteins between k-means clusters

|  | **Protein** | **Protein ID** | **Beta coefficient (standard error)** | **FDR** |
| --- | --- | --- | --- | --- |
| **ULE vs. RRS** | ATP synthase peripheral stalk subunit OSCP | P48047 | 0.33 (0.07) | $6.3 \times 10^{-3}$ |
| | Glucokinase regulatory protein | Q14397 | 0.14 (0.04) | $4.2 \times 10^{-2}$ |
| | Serine/threonine-protein kinase MRCK beta | Q9Y5S2 | 0.09 (0.02) | $4.2 \times 10^{-2}$ |
| | Serine/threonine-protein kinase PAK 6 | Q9NQU5 | 0.37 (0.10) | $4.6 \times 10^{-2}$ |
| | Membrane frizzled-related protein | Q9BY79 | 0.14 (0.04) | $5.5 \times 10^{-2}$ |
| **AD vs. RRS** | Fatty acid-binding protein, heart | P05413 | 0.20 (0.03) | $1.3 \times 10^{-6}$ |
| | Leptin | P41159 | 0.34 (0.06) | $4.8 \times 10^{-6}$ |
| | Renin | P00797 | 0.25 (0.05) | $1.6 \times 10^{-4}$ |
| | Retinoic acid receptor responder protein 2 | Q99969 | 0.08 (0.02) | $2.6 \times 10^{-4}$ |
| | Apolipoprotein M | O95445 | -0.15 (0.03) | $3.9 \times 10^{-4}$ |
| **SE vs. RRS** | Bactericidal permeability-increasing protein | P17213 | 0.40 (0.06) | $1.8 \times 10^{-11}$ |
| | Complement component C9 | P02748 | 0.15 (0.02) | $1.9 \times 10^{-11}$ |
| | Troponin T, cardiac muscle | P45379 | 0.20 (0.03) | $2.1 \times 10^{-10}$ |
| | Protein S100-A12 | P80511 | 0.19 (0.03) | $3.4 \times 10^{-10}$ |
| | Oxidized low-density lipoprotein receptor 1 | P78380 | 0.23 (0.04) | $1.1 \times 10^{-9}$ |
| SOMAscan plasma proteins significantly associated to visit 1 k-means cluster membership from multivariable linear regression modeling (*FDR 10%*). We used linear regression and adjusted for age, sex, race, and current smoking status. The top 5 significantly differentially associated proteins unique to each of the 3 comparisons (ULE vs. RRS, AD vs. RRS, and SE vs. RRS) are reported in this table. The cluster following the “vs.” is the reference group. The units of all proteins are relative fluorescence units. Abbreviations: RRS = Relatively resistant smokers; ULE = Upper lobe predominant emphysema; AD = Airway-predominant disease; SE = Severe emphysema. |  |  |  |  |

Using blood RNA sequencing data from 2,072 subjects at Visit 2, we identified significant associations to 3, 2,105, and 148 genes for the ULE, AD, and SE clusters, respectively with the RRS group as the reference. The Bland-Altman plots are shown in Figure 4 and the complete set of association results is reported in Table S8. Compared to the RRS cluster, the ULE cluster was associated with up-regulation of the *GPR15, AHRR* and *GPR55* genes, and GO pathway enrichment analysis results did not identify any significantly enriched pathways (Table 3). The AD cluster had particularly strong differences in gene expression with 22 and 34 enriched pathways relative to the RRS and SE clusters, respectively. Compared to the RRS cluster, the AD cluster showed significant associations to pathways involved in innate immunity, cellular defense response and NF-kB signaling. Relative to the SE cluster, the AD cluster demonstrated associations to processes involved in both innate and adaptive immunity *(adjusted P-values < 0*.*005)*. The SE cluster also had many differentially expressed genes, with significant pathway enrichment for positive regulation of synapse assembly and cell adhesion. The complete set of pathway enrichment results is reported in Table S9.

**Figure 4.**
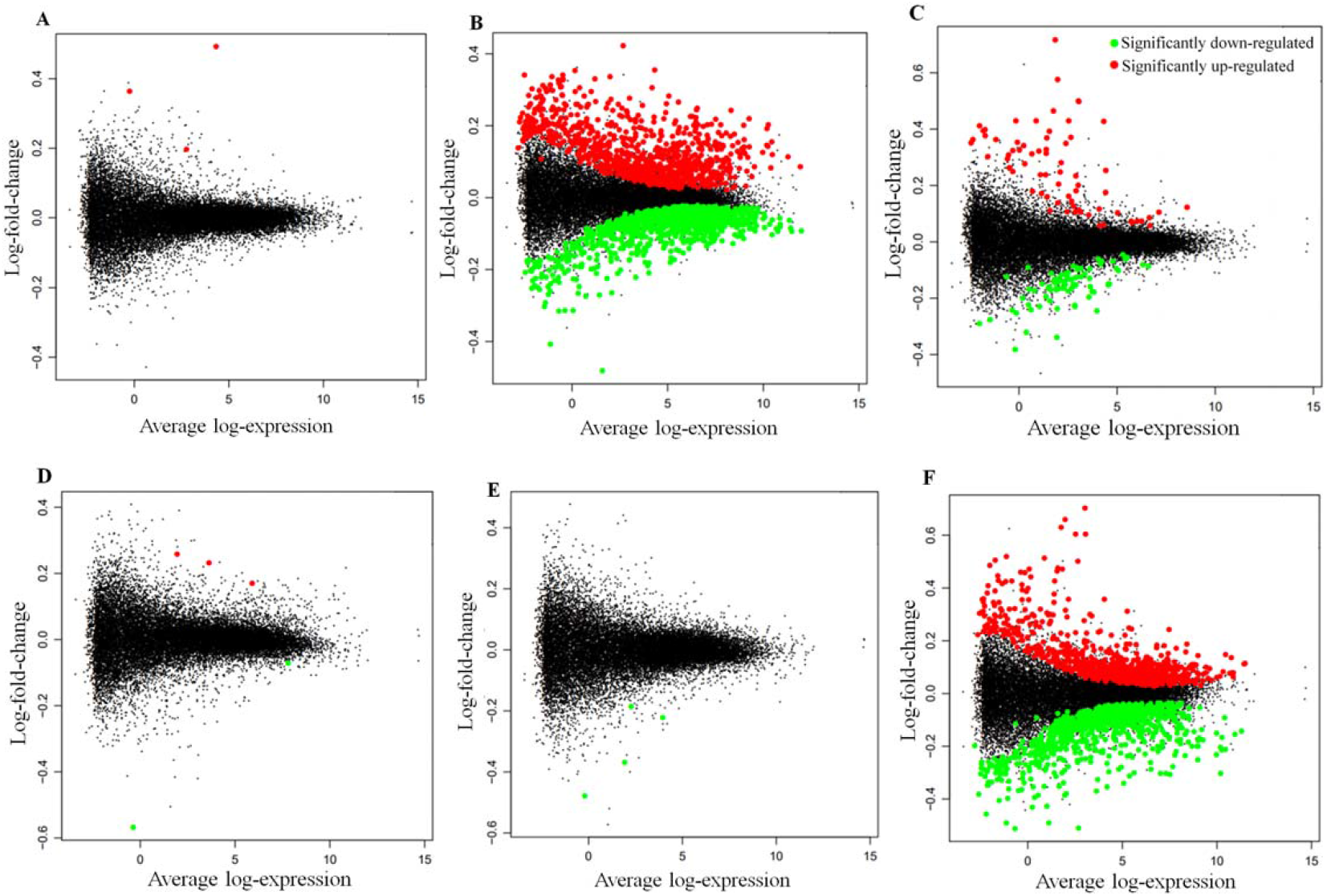
Bland-Altman (MA) plots of the log ratio *versus* mean gene expression for the differential expression analysis results between k-means clusters. The cluster following the “*vs*.” is the reference group. *(A)* ULE *vs*. RRS. *(B)* AD *vs*. RRS. *(C)* SE *vs*. RRS. *(D)* AD *vs*. ULE. *(E)* SE *vs*. ULE. *(F)* SE *vs*. AD. Abbreviations: RRS = Relatively resistant smokers; ULE = Upper lobe predominant emphysema; AD = Airway-predominant disease; SE = Severe emphysema.

**Table 3.**
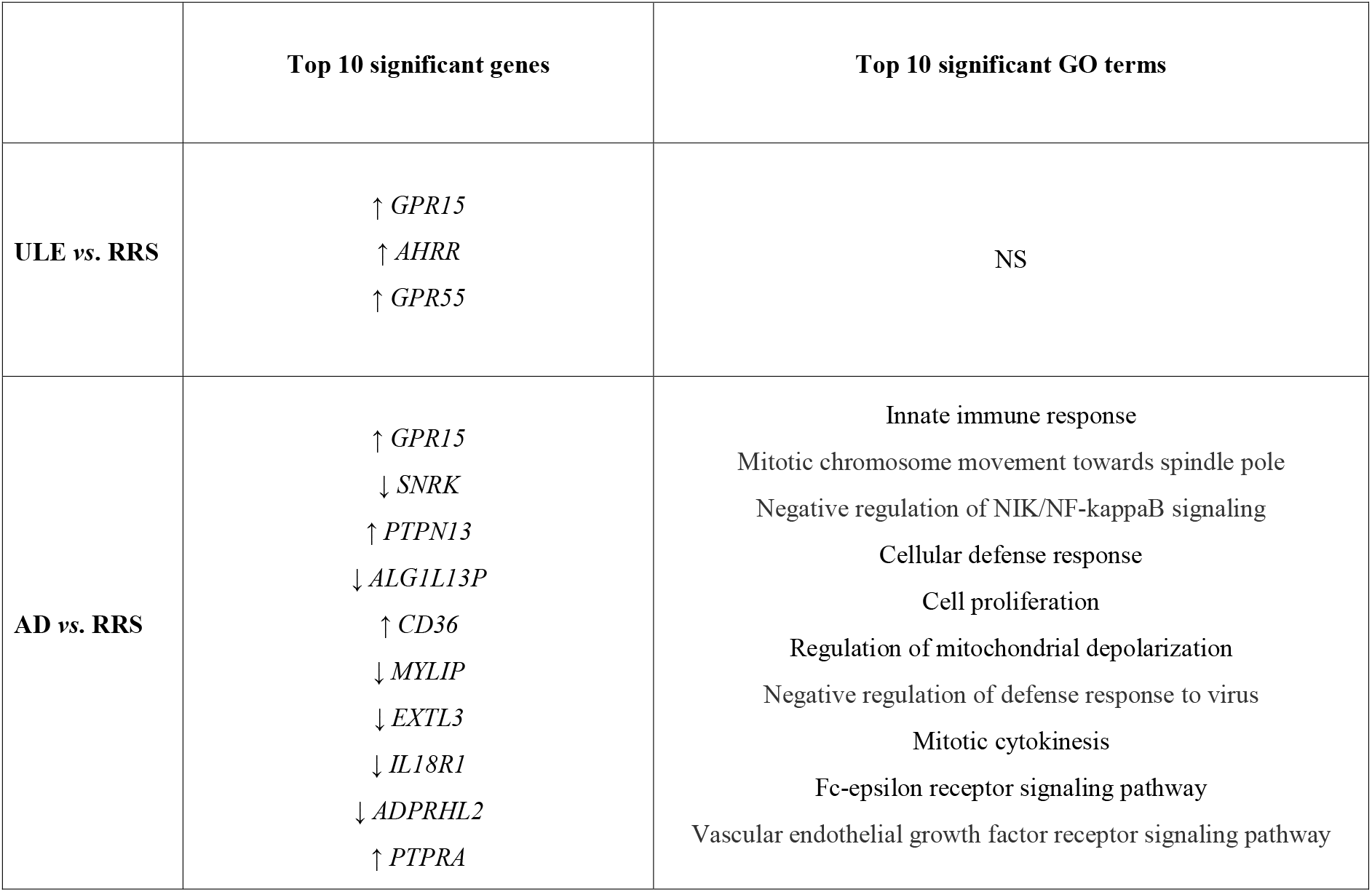
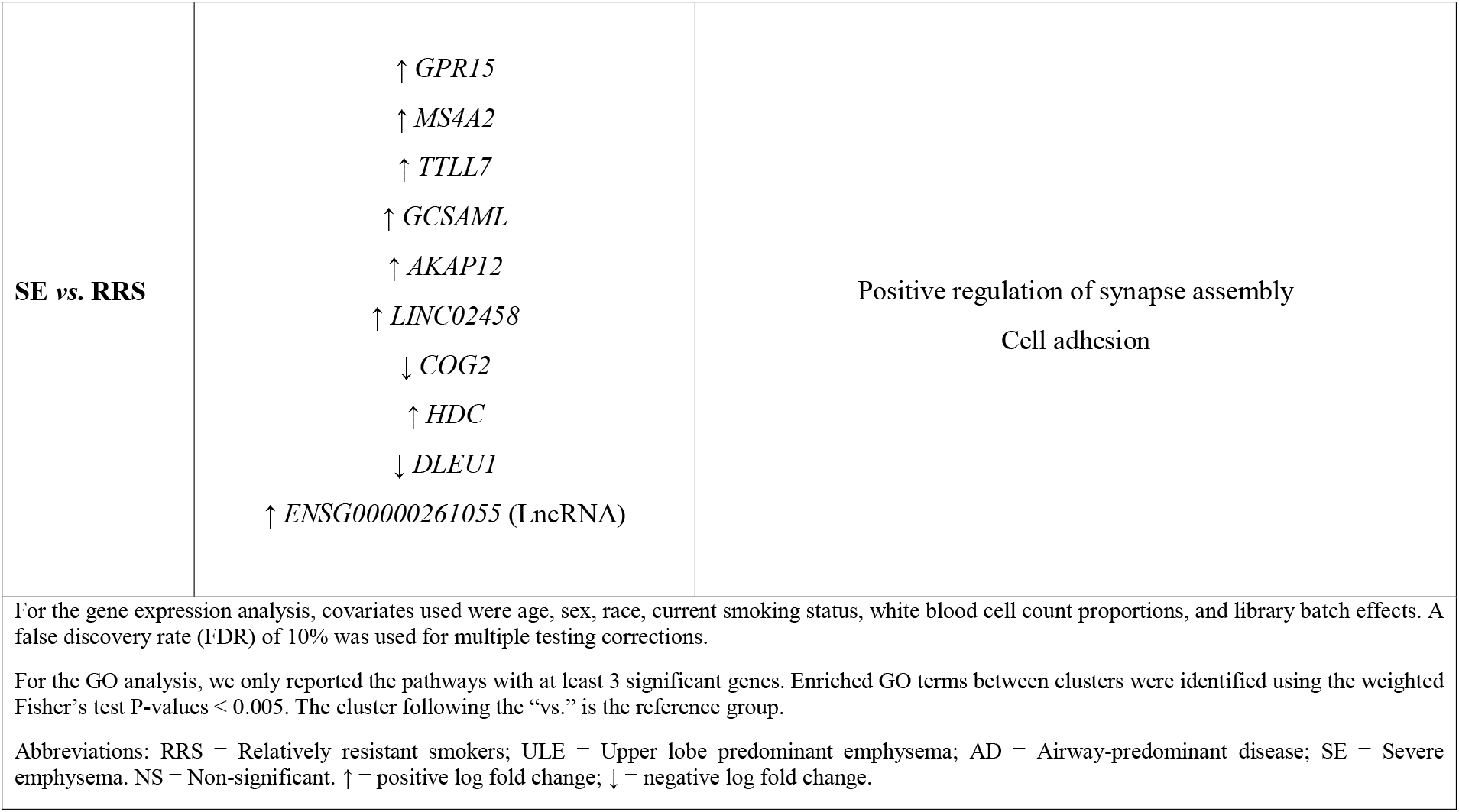
Top 10 significantly differentially expressed genes and enriched gene ontology (GO) terms between k-means clusters

|  | Top 10 significant genes | Top 10 significant GO terms |
| --- | --- | --- |
| **ULE vs. RRS** | ↑ *GPR15*<br>↑ *AHRR*<br>↑ *GPR55* | NS |
| **AD vs. RRS** | ↑ *GPR15*<br>↓ *SNRK*<br>↑ *PTPN13*<br>↓ *ALG1L13P*<br>↑ *CD36*<br>↓ *MYLIP*<br>↓ *EXTL3*<br>↓ *IL18R1*<br>↓ *ADPRHL2*<br>↑ *PTPRA* | Innate immune response<br>Mitotic chromosome movement towards spindle pole<br>Negative regulation of NIK/NF-kappaB signaling<br>Cellular defense response<br>Cell proliferation<br>Regulation of mitochondrial depolarization<br>Negative regulation of defense response to virus<br>Mitotic cytokinesis<br>Fc-epsilon receptor signaling pathway<br>Vascular endothelial growth factor receptor signaling pathway |
| **SE vs. RRS** | ↑ *GPR15*<br>↑ *MS4A2*<br>↑ *TTLL7*<br>↑ *GCSAML*<br>↑ *AKAP12*<br>↑ *LINC02458*<br>↓ *COG2*<br>↑ *HDC*<br>↓ *DLEU1*<br>↑ *ENSG00000261055* (LncRNA) | Positive regulation of synapse assembly<br>Cell adhesion |
| For the gene expression analysis, covariates used were age, sex, race, current smoking status, white blood cell count proportions, and library batch effects. A false discovery rate (FDR) of 10% was used for multiple testing corrections.<br>For the GO analysis, we only reported the pathways with at least 3 significant genes. Enriched GO terms between clusters were identified using the weighted Fisher's test P-values < 0.005. The cluster following the "vs." is the reference group.<br>Abbreviations: RRS = Relatively resistant smokers; ULE = Upper lobe predominant emphysema; AD = Airway-predominant disease; SE = Severe emphysema. NS = Non-significant. ↑ = positive log fold change; ↓ = negative log fold change. |  |  |

## Discussion

In this study, we demonstrated that k-means subgroups of smokers enriched for COPD have varying disease progression patterns, development of prospective comorbidities, and distinct associations to plasma protein and blood transcriptomic biomarkers. The two clusters at the extremes of the lung health spectrum (the RRS and SE clusters) showed high cluster assignment stability between the two visits. Spirometric progression was sensitive to the progression metric being used (absolute *vs*. relative) with the SE subtype showing the most rapid rate of FEV_1_ decline relative to baseline FEV_1_ level. Emphysema progression however was less sensitive to the use of absolute versus relative metrics of progression. In general, the SE cluster had the highest risk for prospective adverse health events, though the AD cluster had the highest risk of incident diabetes and the most distinct gene expression patterns.

A wide array of risk factors have been associated with spirometric decline, such as low BMI, higher baseline FEV_1_ and FVC, smoking exposure, bronchodilator reversibility, African-American race, female sex, previous history of exacerbations, CT-quantified emphysema, upper lobe predominant emphysema and small airway abnormalities ^1 2 30-34^. Whereas previous studies have shown that mild-to-moderate COPD (GOLD 1-2) is associated with an increased loss in absolute FEV_1_ ^2 30^, our study also investigated changes in FEV_1_ relative to baseline values, and found that the SE cluster, which represents a subset of subjects with advanced disease, is associated with more rapid relative FEV_1_ decline. To our knowledge, such differences in the metrics of progression used to assess COPD progression have not been previously reported. This finding emphasizes the fact that disease activity and disease severity are distinct concepts that should be considered when assessing COPD patients ^1 35^.

In contrast to FEV_1_ changes, less is known about the factors that are associated with emphysema progression ^36-38^. In our study, we evaluated emphysema changes by COPD subtype, investigated both absolute and relative changes, and adjusted the analysis for age, sex, race, BMI, smoking pack-years, and CT scanner types. We showed that relative changes were the highest in the SE cluster, which has more advanced baseline emphysema, low BMI, and more COPD exacerbations.

In regard to prospective COPD events, prior reports have indicated that more severe airflow obstruction and COPD exacerbation history are associated with higher risks of COPD exacerbations and CVD ^7 39-41^. We demonstrated that, when adjusted for covariates including airflow limitation and COPD exacerbation history, the SE cluster had a 3 times higher risk of a COPD exacerbation, while the AD and ULE clusters had a hazard ratio of ∼1.35 relative to the RRS group. The SE cluster was also associated with the highest risk of incident CVD, which may be explained by atherosclerosis or arterial stiffness mediated by inflammatory markers ^42^. Another novel finding from our study was that the AD cluster had the highest risk for the prospective development of diabetes, even after adjusting for BMI. This adds to the finding from the study by Hersh et al., which revealed that COPD with limited emphysema and high airflow obstruction is associated with diabetes in cross-sectional data ^43^. Our study provides further evidence linking airway-predominant COPD to diabetes and metabolic syndrome.

The age, dyspnea, and airflow obstruction (ADO) and BODE indices have been classically used to predict mortality in COPD, but less is known about the COPD characteristics that contribute to cause-specific mortality ^44 45^. CVD, airflow obstruction, low BMI, emphysema, and poor exercise capacity were found to be associated with a high risk of all-cause mortality in COPD ^39 46-48^. We similarly observed that the SE cluster, which had these risk factors at baseline, had the highest risk of all-cause mortality. With regards to the CVD mortality, high MMRC dyspnea score, exacerbation history, low FEV_1_, and accelerated lung function decline have been shown to be contributors ^49 50^. In our paper, we showed that the SE, AD, and ULE clusters had elevated risks of CVD-related mortality in unadjusted models, but when correcting for age, sex, race, BODE, and smoking pack-years, only the AD and ULE clusters had significant risks, which indicates that airway disease and lobular emphysema distribution may be additional contributors to this risk. While most previously published COPD studies have examined the association with prospective cancer rather than prospective mortality due to cancer, some reports have indicated that COPD is associated with an elevated risk of both lung and extra-pulmonary cancer mortalities ^51 52^. In our study, similar to CVD mortality, the ULE, AD, and SE clusters had elevated cancer mortality risks, but after adjustments, only the ULE cluster had a significantly increased cancer mortality risk. Differential genetic susceptibility, delayed clearance of inhaled carcinogens, and chronic inflammation may be potential mechanisms underlying these associations ^52^.

COPD subtype-specific associations to protein and transcriptomic biomarkers have only been studied in a few studies of relatively small size ^14^. In our analysis, the AD cluster had elevated plasma levels of leptin, supporting a previous report which showed that pro-inflammatory adipokines leptin and adiponectin are implicated in COPD ^53^. While many previous studies have shown that innate immunity is related to COPD ^54 55^, we observed that it was the SE cluster that demonstrated the strongest association to proteins involved in the innate immune response. At the transcriptomic level, COPD has been associated with inflammation and sphingolipid metabolism ^15-17^, and our study demonstrates that it is the AD subtype that has the most distinct gene expression signature that is enriched for these pathways.

This study has a number of strengths. Compared to previous publications, our study included a larger sample size and longer follow-up, and investigated incident comorbidities in addition to lung health outcomes. Additionally, we included a well-phenotyped cohort of smokers across the full spectrum of disease severity, and we were able to test the association of subtypes to clinical, radiologic, and multi-omic molecular markers. When studying progression, we considered both absolute and relative changes in lung function and CT-quantified emphysema. Furthermore, this is to our knowledge the first study demonstrating that airway-predominant COPD is independently associated with incident diabetes risk and has particularly strong associations to inflammatory biomarkers.

One of the limitations of our study is that because of its observational design, the statistical associations observed may not reflect causal relationships. Our study sample was limited to subjects who survived the five-year observation period and as a result, our findings are not representative of subjects with very advanced COPD and limited life expectancy. Analyses that jointly model death and other aspects of disease progression would provide additional useful information. As further follow-up data from COPDGene is obtained, our hypotheses from this study can be validated over longer follow-up.

## Conclusions

COPD-related subtypes defined by spirometric and radiologic measures at baseline have different rates of disease progression and are differentially associated to prospective health outcomes. They also exhibit distinct biomarker profiles indicative of underlying biological differences. In the future, these subtypes could be used as the basis for targeted drug development, studies of differential treatment response, or the enrollment of specific subgroups in clinical trials.

## Data Availability

All data produced in the present study are available upon reasonable request to the authors.

## ACKNOWLEDGEMENTS

**COPDGene Investigators -Core Units:**

*Administrative Center*: James D. Crapo, MD (PI); Edwin K. Silverman, MD, PhD (PI); Barry J. Make, MD; Elizabeth A. Regan, MD, PhD

*Genetic Analysis Center*: Terri H. Beaty, PhD; Peter J. Castaldi, MD, MSc; Michael H. Cho, MD, MPH; Dawn L. DeMeo, MD, MPH; Adel Boueiz, MD, MMSc; Marilyn G. Foreman, MD, MS; Auyon Ghosh, MD; Lystra P. Hayden, MD, MMSc; Craig P. Hersh, MD, MPH; Jacqueline Hetmanski, MS; Brian D. Hobbs, MD, MMSc; John E. Hokanson, MPH, PhD; Wonji Kim, PhD; Nan Laird, PhD; Christoph Lange, PhD; Sharon M. Lutz, PhD; Merry-Lynn McDonald, PhD; Dmitry Prokopenko, PhD; Matthew Moll, MD, MPH; Jarrett Morrow, PhD; Dandi Qiao, PhD; Elizabeth A. Regan, MD, PhD; Aabida Saferali, PhD; Phuwanat Sakornsakolpat, MD; Edwin K. Silverman, MD, PhD; Emily S. Wan, MD; Jeong Yun, MD, MPH

*Imaging Center*: Juan Pablo Centeno; Jean-Paul Charbonnier, PhD; Harvey O. Coxson, PhD; Craig J. Galban, PhD; MeiLan K. Han, MD, MS; Eric A. Hoffman, Stephen Humphries, PhD; Francine L. Jacobson, MD, MPH; Philip F. Judy, PhD; Ella A. Kazerooni, MD; Alex Kluiber; David A. Lynch, MB; Pietro Nardelli, PhD; John D. Newell, Jr., MD; Aleena Notary; Andrea Oh, MD; Elizabeth A. Regan, MD, PhD; James C. Ross, PhD; Raul San Jose Estepar, PhD; Joyce Schroeder, MD; Jered Sieren; Berend C. Stoel, PhD; Juerg Tschirren, PhD; Edwin Van Beek, MD, PhD; Bram van Ginneken, PhD; Eva van Rikxoort, PhD; Gonzalo Vegas SanchezFerrero, PhD; Lucas Veitel; George R. Washko, MD; Carla G. Wilson, MS

*PFT QA Center, Salt Lake City, UT*: Robert Jensen, PhD

*Data Coordinating Center and Biostatistics, National Jewish Health, Denver, CO*: Douglas Everett, PhD; Jim Crooks, PhD; Katherine Pratte, PhD; Matt Strand, PhD; Carla G. Wilson, MS

*Epidemiology Core, University of Colorado Anschutz Medical Campus, Aurora*, CO: John E. Hokanson, MPH, PhD; Erin Austin, PhD; Gregory Kinney, MPH, PhD; Sharon M. Lutz, PhD; Kendra A. Young, PhDVersion Date: March 26, 2021

*Mortality Adjudication Core*: Surya P. Bhatt, MD; Jessica Bon, MD; Alejandro A. Diaz, MD, MPH; MeiLan K. Han, MD, MS; Barry Make, MD; Susan Murray, ScD; Elizabeth Regan, MD; Xavier Soler, MD; Carla G. Wilson, MS

*Biomarker Core*: Russell P. Bowler, MD, PhD; Katerina Kechris, PhD; Farnoush BanaeiKashani, PhD

**COPDGene Investigators -Clinical Centers:**

*Ann Arbor VA*: Jeffrey L. Curtis, MD; Perry G. Pernicano, MD

*Baylor College of Medicine, Houston, TX*: Nicola Hanania, MD, MS; Mustafa Atik, MD; Aladin Boriek, PhD; Kalpatha Guntupalli, MD; Elizabeth Guy, MD; Amit Parulekar, MD

*Brigham and Women’s Hospital, Boston, MA*: Dawn L. DeMeo, MD, MPH; Craig Hersh, MD, MPH; Francine L. Jacobson, MD, MPH; George Washko, MD

*Columbia University, New York, NY*: R. Graham Barr, MD, DrPH; John Austin, MD; Belinda D’Souza, MD; Byron Thomashow, MD

*Duke University Medical Center, Durham, NC*: Neil MacIntyre, Jr., MD; H. Page McAdams, MD; Lacey Washington, MD

*HealthPartners Research Institute*, Minneapolis, MN: Charlene McEvoy, MD, MPH; Joseph Tashjian, MD

*Johns Hopkins University*, Baltimore, MD: Robert Wise, MD; Robert Brown, MD; Nadia N. Hansel, MD, MPH; Karen Horton, MD; Allison Lambert, MD, MHS; Nirupama Putcha, MD, MHS

*Lundquist Institute for Biomedical Innovation at Harbor UCLA Medical Center*, Torrance, CA: Richard Casaburi, PhD, MD; Alessandra Adami, PhD; Matthew Budoff, MD; Hans Fischer, MD; Janos Porszasz, MD, PhD; Harry Rossiter, PhD; William Stringer, MD

*Michael E. DeBakey VAMC*, Houston, TX: Amir Sharafkhaneh, MD, PhD; Charlie Lan, DO

*Minneapolis VA*: Christine Wendt, MD; Brian Bell, MD; Ken M. Kunisaki, MD, MS

*Morehouse School of Medicine, Atlanta, GA*: Eric L. Flenaugh, MD; Hirut Gebrekristos, PhD; Mario Ponce, MD; Silanath Terpenning, MD; Gloria Westney, MD, MS

*National Jewish Health, Denver, CO*: Russell Bowler, MD, PhD; David A. Lynch, MB Reliant Medical Group, Worcester, MA: Richard Rosiello, MD; David Pace, MD

*Temple University, Philadelphia, PA*: Gerard Criner, MD; David Ciccolella, MD; Francis Cordova, MD; Chandra Dass, MD; Gilbert D’Alonzo, DO; Parag Desai, MD; Michael Jacobs, PharmD; Steven Kelsen, MD, PhD; Victor Kim, MD; A. James Mamary, MD; Nathaniel Marchetti, DO; Aditi Satti, MD; Kartik Shenoy, MD; Robert M. Steiner, MD; Alex Swift, MD; Irene Swift, MD; Maria Elena Vega-Sanchez, MD

*University of Alabama, Birmingham, AL*: Mark Dransfield, MD; William Bailey, MD; Surya P. Bhatt, MD; Anand Iyer, MD; Hrudaya Nath, MD; J. Michael Wells, MD

*University of California, San Diego, CA*: Douglas Conrad, MD; Xavier Soler, MD, PhD; Andrew Yen, MD

*University of Iowa, Iowa City, IA*: Alejandro P. Comellas, MD; Karin F. Hoth, PhD; John Newell, Jr., MD; Brad Thompson, MD

*University of Michigan, Ann Arbor, MI*: MeiLan K. Han, MD MS; Ella Kazerooni, MD MS; Wassim Labaki, MD MS; Craig Galban, PhD; Dharshan Vummidi, MD

*University of Minnesota, Minneapolis, MN*: Joanne Billings, MD; Abbie Begnaud, MD; Tadashi Allen, MD

*University of Pittsburgh, Pittsburgh, PA*: Frank Sciurba, MD; Jessica Bon, MD; Divay Chandra, MD, MSc; Joel Weissfeld, MD, MPH

*University of Texas Health, San Antonio, San Antonio, TX*: Antonio Anzueto, MD; Sandra Adams, MD; Diego Maselli-Caceres, MD; Mario E. Ruiz, MD; Harjinder Singh

