## Supplementary Materials for "Clustering-based COPD Subtypes Have Distinct Longitudinal Outcomes and Multi-omics Biomarkers"

Andrew Gregory^1^, Zhonghui Xu^1^, Katherine Pratte^2^, Sool Lee^1^, Congjian Liu^3^, Robert Chase^1^, Jeong H. Yun^1,3^, Aabida Saferali^1^, Craig P. Hersh^1,3^, Russell P. Bowler^4^, Edwin K. Silverman^1,3^, Peter J. Castaldi^1,5^*, Adel Boueiz^1,3^* for the COPDGene investigators.

*Contributed equally

^1^Channing Division of Network Medicine, Brigham and Women’s Hospital, Harvard Medical School, Boston, MA; ^2^Department of Biostatistics, National Jewish Health, Denver, CO; ^3^Pulmonary and Critical Care Medicine, Brigham and Women’s Hospital, Harvard Medical School, Boston, MA; ^4^Division of Pulmonary, Critical Care and Sleep Medicine, National Jewish Health, Denver, Colorado; ^5^General Medicine and Primary Care, Brigham and Women’s Hospital, Harvard Medical School, Boston, MA

**Methods**

***COPDGene***

Exclusion criteria of subjects in the COPDGene study included a history of other lung diseases except asthma, prior lobectomy or lung volume reduction surgery, active cancer undergoing treatment, known or suspected lung cancer, or pregnancy. A severe exacerbation was defined as a COPD exacerbation that required a hospital admission or an emergency room visit. Quality of life and respiratory disease-related health impairments were determined using the St George’s Respiratory Questionnaire (SGRQ) ^1^. Dyspnea was assessed by the Modified Medical Research Council (MMRC) dyspnea score system ^2^. Metabolic syndrome was defined as having at least 3 of the following conditions: BMI > 30, diabetes, hypertension, and high cholesterol. During both visits in the COPDGene study, spirometry was performed before and after 180 mcg of albuterol was administered (ndd Easy-One spirometer, Andover, MA). The Hankinson NHANES reference equations were used to calculate the percent predicted values ^3^. Normal spirometry (GOLD grade 0) was defined as a post-bronchodilator FEV_1_/FVC ≥ 0.7 and FEV_1_ ≥ 80% predicted. GOLD 1-4 were defined as FEV_1_/FVC < 0.70 and post-bronchodilator FEV_1_ ≥ 80% predicted (GOLD 1), FEV_1_/FVC < 0.70 and post-bronchodilator FEV_1_ 50-79% predicted (GOLD 2), FEV_1_/FVC < 0.70 and post-bronchodilator FEV_1_ 30-49% predicted (GOLD 3), and FEV_1_/FVC < 0.70 and post-bronchodilator FEV_1_ < 30% predicted (GOLD 4) ^4^. More information about the COPDGene is available elsewhere ^5^.

***Radiologic assessment***

The log of the lung upper third to lower third ratio of emphysema (%LAA-950) was used to evaluate the distribution of apico-basal emphysema. VIDA software ([www.vidadiagnostics.com](about:blank)) measured airway disease as gas trapping (percentage of low attenuation units < -856 HU at end-expiration), airway wall thickness (obtained along the center line of the lumen, in the middle third of the airway segment, for one segmental airway of each lung lobe; the mean value across all lobes was used for analysis), and Pi10 (the square root of the wall area of a hypothetical airway of 10-mm internal perimeter).

***Plasma protein biomarkers***

At baseline, participants who agreed and consented to participate in an omics ancillary study provided an additional sample of blood, collected using 8.5 mL p100 tubes (Becton Dickinson and Company). National Jewish Health stored and determined protein levels of 1,305 proteins using the SOMAscan Human Plasma 1.3K assay (SomaLogic, Boulder, Colorado). SOMAscan is a multiplex aptamer-based assay. Aptamers are single-stranded deoxyoligonucleotides that bind with high affinity and specificity to specific protein structures ^6^. SOMAscan data was standardized by SomaLogic per their protocol. It consists of within plate hybridization to control for variability across array signals, median signal normalization to control for technical variability of replicates within a run, and plate scaling and calibration of SOMAmers to control for inter-assay variation between analytes and batch differences between plates.

***Total RNA extraction***

Total blood RNA was extracted from subjects at Visit 2 and collected in PAXgene TM Blood RNA tubes from the Qiagen PreAnalytiX PAXgene Blood miRNA Kit (Qiagen, Valencia, CA). The extraction protocol was performed either with the Qiagen QIAcube extraction robot according to the company’s standard operating procedure or manually. RNA samples with a concentration of ≥ 25 μg/ul and RNA integrity number (RIN) > 6 were sequenced.

***cDNA library construction and sequencing***

Total RNA Globin reduction and cDNA library preparation were performed with the Illumina TruSeq Stranded Total RNA with Ribo-Zero Globin kit (Illumina, Inc., San Diego, CA). Quantification with picogreen, size analysis on an Agilent Bioanalyzer or Tapestation 2200 (Agilent, Santa Clara, CA), and qPCR quantitation against a standard curve assisted with library quantity control. Samples were sequenced to an average depth of 20 million 75bp paired end reads on Ilumina HiSeq 2500 sequencers.

***Sequencing read alignment, quality control and expression quantification***

Skewer with default parameters ^7^ trimmed reads of TruSeq adapters. The STAR (version 2.5.2b) aligner ^8^ aligned the trimmed reads to the GRCh38 genome. Transcript GTF and gene annotations were downloaded from the Biomart Ensembl database (Ensembl Genes release 94, GRCh38.p12 assembly). Quality control was performed with the FastQC ^9^ and RNA-SeQC programs ^10^ and samples were included for further analysis if they had > 10 million total reads, > 80% of reads mapped to the reference genome, XIST and Y chromosome expression was consistent with reported sex, < 10% of R1 reads in the sense orientation, Pearson correlation ≥ 0.9 with samples in the same library construction batch, and concordant genotype calls between variants called from RNA sequencing reads and DNA genotyping. Sequencing read counts were obtained from the featureCounts function in the Rsubread R package (v1.32.2) and the gene count data used for this analysis are available in GEO ^11 12^ (accession number GSE158699).

***Differential gene expression***

Genes were filtered to remove very low expressed genes (average counts per million (CPM) < 0.2 or number of subjects with CPM < 0.5 was < 50) or outlying extremely highly expressed genes (the number of subjects with CPM > 50,000 was less than 50). The trimmed mean of M values (TMM) procedure from the edgeR R package (v3.24.3) was applied to account for differences in sequencing depth. Subsequently, counts were transformed to log2 CPM values and quantile-normalized to further remove systematic noise from the data.

**Figure Legends:**

**Figure S1:** Study flow chart. Abbreviations: NHW = non-Hispanic whites. AA = African-Americans.

**Figure S2:** Changes in cluster assignments at Visit 2. Percentage of subjects in each cluster at Visit 2. Abbreviations: RRS = Relatively resistant smokers; ULE = Upper lobe predominant emphysema; AD = Airway-predominant disease; SE = Severe emphysema.

**Figure S3:** Kaplan-Meier plots of mortality by k-means cluster. *(A)* Risk of a respiratory-related mortality. *(B)* Risk of CVD-related mortality. *(C)* Risk of cancer-related mortality. *(D)* Risk of mortality due to other causes. Limited data is available for subjects at the 8-year time point, since the mortality adjudication process is still ongoing. Abbreviations: RRS = Relatively resistant smokers; ULE = Upper lobe predominant emphysema; AD = Airway-predominant disease; SE = Severe emphysema.

**Figure S4:** UpSet plots of SOMAscan plasma proteins significantly associated with k-means cluster membership. Reference group was the RRS cluster. Covariates used were age, sex, race, and current smoking status. We corrected for multiple comparisons with the Benjamini-Hochberg method.  Proteins were selected if they reached a false discovery rate (FDR) of 10%. Abbreviations: RRS = Relatively resistant smokers; ULE = Upper lobe predominant emphysema; AD = Airway-predominant disease; SE = Severe emphysema.

**Table S1.** Visit 1 characteristics of subjects who were included *vs*. excluded in the analyses of longitudinal changes in FEV_1_ and emphysema.

|  | **Included subjects**  **(n = 4,679)** | **Excluded subjects**  **(n = 3,587)** | **P-value** |
| --- | --- | --- | --- |
| Age | 59.60 (13.70) | 57.90 (15.30) | *< 0.0001* |
| Sex, % male | 51.10 | 57.04 | *< 0.0001* |
| BMI | 28.25 (7.61) | 27.71 (8.09) | *< 0.0001* |
| Smoking pack-years | 38.70 (27.30) | 40.40 (27.70) | *< 0.0001* |
| FEV_1_ (mL) | 2,326 (1,178) | 2,173.50 (1,470) | *< 0.0001* |
| FEV_1_, % predicted | 83.50 (29.95) | 76.90 (40.70) | *< 0.0001* |
| FVC (mL) | 3,299 (1,368) | 3,159 (1,436) | *< 0.0001* |
| GOLD  PRISm  GOLD 0  GOLD 1  GOLD 2  GOLD 3  GOLD 4 | 558 (54.81 %)  2,220 (61.26 %)  419 (64.07 %)  918 (57.3 %)  466 (50.27 %)  98 (22.37 %) | 460 (45.19 %)  1,404 (38.74 %)  235 (35.93 %)  684 (42.70 %)  461 (49.73 %)  340 (77.63 %) | 0.2  *< 0.0001*  *< 0.0001*  0.5  *< 0.0001*  *< 0.0001* |
| Bronchodilator responsiveness (% FEV_1_) | 4.29 (8.47) | 4.55 (9.65) | 0.2 |
| Bronchodilator responsiveness (% FVC) | 1.86 (10.02) | 2.46 (11.36) | *0.03* |
| Adjusted Perc15 density | 86.08 (29.91) | 85.59 (33.89) | *0.01* |
| %LAA-950 | 1.91 (5.39) | 2.04 (7.86) | *0.02* |
| Upper/lower emphysema ratio | 1.00 (1.55) | 1.20 (1.74) | *< 0.0001* |
| % Segmental airway wall thickness | 49.12 (11.57) | 52.27 (12.22) | *< 0.0001* |
| Gas trapping (%) | 13.54 (20.77) | 14.91 (29.97) | *< 0.0001* |
| Pi10 | 2.15 (0.75) | 2.38 (0.89) | *< 0.0001* |
| Exacerbation history (%) | 17.97 | 23.98 | *< 0.0001* |
| Severe exacerbations history (%) | 8.21 | 15.25 | *< 0.0001* |
| SGRQ symptom score | 23.39 (38.24) | 34.11 (43.29) | *< 0.0001* |
| MMRC dyspnea score  0  1  2  3  4 | 2,346 (63.27 %)  717 (60.4 %)  604 (55.72 %)  710 (47.24 %)  302 (38.52 %) | 1,362 (36.73 %)  470 (39.6 %)  480 (44.28 %)  793 (52.76 %)  482 (61.48 %) | *< 0.0001*  *0.004*  0.5  *< 0.0001*  *< 0.0001* |
| CVD (%) | 16.48 % | 17.74 % | 0.1 |
| Diabetes (%) | 12.44 % | 13.49 % | 0.2 |
| Hypertension (%) | 42.45 % | 44.24 % | 0.1 |
| Mortality, n | 0 | 935 | *< 0.0001* |
| Continuous variables are reported as medians (interquartile ranges). Categorical variables are reported as percentages. Kruskal-Wallis rank sum tests were used for continuous variables. Chi-square tests were used for categorical variables.  BMI: Body mass index; Bronchodilator responsiveness (%) FEV_1_: Percentage of subjects with post-bronchodilator increase in FEV_1_ of at least 12% from baseline; Bronchodilator responsiveness (%) FVC: Percentage of subjects with post-bronchodilator increase in FVC of at least 12% from baseline; CVD: Cardiovascular disease (composite endpoint of stroke, heart attack, coronary artery disease, coronary artery bypass graft surgery, peripheral artery disease, and/or cardiac angina); Exacerbation history: At least one COPD exacerbation (acute worsening of respiratory symptoms that required systemic steroids and/or antibiotics) in the previous year; Severe exacerbation history: COPD exacerbation requiring an emergency department visit or hospital admission; FEV_1_ (mL): Forced expiratory volume in 1 second; FEV_1_, % predicted: Percent of the normal FEV_1_ based on height, weight, and race; FVC: Forced vital capacity; GOLD: Global Initiative for Chronic Obstructive Lung Disease; GOLD 0: Normal spirometry (defined as post-bronchodilator FEV_1_/FVC ≥ 0.7 and FEV_1_ ≥ 80% predicted); GOLD 1: FEV_1_/FVC < 0.70 and post-bronchodilator FEV_1_ ≥ 80% predicted; GOLD 2: FEV_1_/FVC < 0.70 and post-bronchodilator FEV_1_ 50-79% predicted; GOLD 3: FEV_1_/FVC < 0.70 and post-bronchodilator FEV_1_ 30-49% predicted; GOLD 4: FEV_1_/FVC < 0.70 and post-bronchodilator FEV_1_ < 30% predicted; MMRC: Modified medical research council dyspnea scoring system; PRISm: Preserved Ratio Impaired Spirometry (defined as FEV_1_/FVC ≥ 0.70 but with FEV_1_ < 80% predicted); SGRQ: St. George’s respiratory questionnaire score.  Adjusted Perc15 density: Cut off value in Hounsfield units (HU) below which 15% of all voxels are distributed on a lung CT scan (per convention, adjusted Perc15 density values are reported as the HU + 1000); Gas trapping (%): Percentage of lung voxels with a density less than -856 HU at end exhalation; % LAA-950: Percentage of CT low attenuation less than -950 HU at end-inspiration using Thirona software; Pi10: Square root of the wall area of a hypothetical airway of a 10-mm internal perimeter; % Segmental airway wall thickness: Percentage of the wall relative to the total bronchial area for the segmental airways; Upper/lower emphysema ratio: Log of the lung upper third to lower third ratio of emphysema.  P-values correspond to global P-values for comparisons across all 4 k-means clusters. P-values < 0.05 are italicized. | | | |

**Table S2.** Visit 1 characteristics of k-means clusters

|  | **Relatively Resistant Smokers**  **(n = 3,192)** | **Mild Upper Lobe Predominant Emphysema**  **(n = 1,242)** | **Airway Predominant Disease**  **(n = 2,176)** | **Severe Emphysema**  **(n = 1,656)** | **P-value** |
| --- | --- | --- | --- | --- | --- |
| Age | 57.90 (13.70) | 57.40 (12.48) | 55.20 (12.65) | 66.00 (11.53) | *< 0.0001* |
| Sex, % male | 56.45 | 48.15 | 47.70 | 60.33 | *< 0.0001* |
| BMI | 27.74 (6.88) | 26.83 (7.39) | 30.91 (8.95) | 26.26 (7.41) | *< 0.0001* |
| Smoking pack-years | 35.00 (24.20) | 41.70 (25.85) | 38.00 (25.80) | 50.00 (34.45) | *< 0.0001* |
| FEV_1_ (mL) | 2,838.50 (1,017.50) | 2,296.00 (963.75) | 2,127.00 (920.50) | 1,094.00 (682.75) | *< 0.0001* |
| FEV_1_, % predicted | 95.30 (18.90) | 81.45 (21.17) | 74.90 (22.50) | 40.50 (22.65) | *< 0.0001* |
| FVC (mL) | 3,720 (1,347) | 3,250 (1,224) | 2,972 (1,206) | 2,681 (1,210) | *< 0.0001* |
| GOLD  PRISm  GOLD 0  GOLD 1  GOLD 2  GOLD 3  GOLD 4 | 224 (22 %)  2,412 (66.56 %)  346 (52.91 %)  204 (12.73 %)  6 (0.65 %)  0 (0 %) | 186 (18.27 %)  487 (13.44 %)  186 (28.44 %)  361 (22.53 %)  21 (2.27 %)  1 (0.23 %) | 596 (58.55 %)  723 (19.95 %)  108 (16.51 %)  572 (35.71 %)  158 (17.04 %)  18 (4.11 %) | 12 (1.18 %)  2 (0.06 %)  14 (2.14 %)  465 (29.03 %)  742 (80.04 %)  419 (95.66 %) | *< 0.0001* |
| Bronchodilator responsiveness (% FEV_1_) | 3.35 (6.43) | 3.89 (8.36) | 4.74 (10.59) | 7.93 (13.53) | *< 0.0001* |
| Bronchodilator responsiveness (% FVC) | 0.51 (7.53) | 1.64 (9.83) | 2.92 (11.86) | 6.58 (14.54) | *< 0.0001* |
| Adjusted Perc15 density | 88.95 (24.48) | 89.77 (27.56) | 96.59 (27.23) | 52.04 (28.38) | *< 0.0001* |
| Emphysema (%LAA-950) | 1.40 (3.08) | 2.10 (3.97) | 0.69 (1.62) | 18.85 (16.83) | *< 0.0001* |
| Upper/lower emphysema ratio | 0.69 (0.47) | 3.87 (6.51) | 0.52 (0.41) | 1.43 (1.73) | *< 0.0001* |
| % Segmental airway wall thickness | 44.73 (7.65) | 50.33 (10.54) | 56.89 (9.78) | 54.54 (10.34) | *< 0.0001* |
| Gas trapping (%) | 9.87 (12.86) | 13.87 (15.63) | 9.91 (13.76) | 52.32 (22.25) | *< 0.0001* |
| Pi10 | 1.86 (0.41) | 2.23 (0.63) | 2.65 (0.76) | 2.7 (0.69) | *< 0.0001* |
| Exacerbation history (%) | 8.77 | 17.39 | 21.65 | 44.32 | *< 0.0001* |
| Severe exacerbations history (%) | 3.26 | 10.55 | 13.28 | 24.58 | *< 0.0001* |
| SGRQ symptom score | 15.01 (28.78) | 28.61 (40.06) | 33.88 (42.96) | 47.68 (35.80) | *< 0.0001* |
| MMRC dyspnea score  0  1  2  3  4 | 2,089 (56.34 %)  466 (39.26 %)  274 (25.28 %)  276 (18.36 %)  87 (11.1 %) | 544 (14.67 %)  211 (17.78 %)  179 (16.51 %)  215 (14.3 %)  93 (11.86 %) | 866 (23.35 %)  319 (26.87 %)  314 (28.97 %)  441 (29.34 %)  236 (30.1 %) | 209 (5.64 %)  191 (16.09 %)  317 (29.24 %)  571 (37.99 %)  368 (46.94 %) | *< 0.0001* |
| CVD (%) | 12.03 % | 17.97 % | 18.53 % | 23.99 % | *< 0.0001* |
| Diabetes (%) | 10.49 % | 10.47 % | 19.44 % | 10.75 % | *< 0.0001* |
| Hypertension (%) | 36.98 % | 41.87 % | 48.25 % | 49.70 % | *< 0.0001* |
| Continuous variables are reported as medians (interquartile ranges). Categorical variables are reported as percentages. Kruskal-Wallis rank sum tests were used for continuous variables. Chi-square tests were used for categorical variables.  BMI: Body mass index; Bronchodilator responsiveness (%) FEV_1_: Percentage of subjects with post-bronchodilator increase in FEV_1_ of at least 12% from baseline; Bronchodilator responsiveness (%) FVC: Percentage of subjects with post-bronchodilator increase in FVC of at least 12% from baseline; CVD: Cardiovascular disease (composite endpoint of stroke, heart attack, coronary artery disease, coronary artery bypass graft surgery, peripheral artery disease, and/or cardiac angina); Exacerbation history: At least one COPD exacerbation (acute worsening of respiratory symptoms that required systemic steroids and/or antibiotics) in the previous year; Severe exacerbation history: COPD exacerbation requiring an emergency department visit or hospital admission; FEV_1_ (mL): Forced expiratory volume in 1 second; FEV_1_, % predicted: Percent of the normal FEV_1_ based on height, weight, and race; FVC: Forced vital capacity; GOLD: Global Initiative for Chronic Obstructive Lung Disease; GOLD 0: Normal spirometry (defined as post-bronchodilator FEV_1_/FVC ≥ 0.7 and FEV_1_ ≥ 80% predicted); GOLD 1: FEV_1_/FVC < 0.70 and post-bronchodilator FEV_1_ ≥ 80% predicted; GOLD 2: FEV_1_/FVC < 0.70 and post-bronchodilator FEV_1_ 50-79% predicted; GOLD 3: FEV_1_/FVC < 0.70 and post-bronchodilator FEV_1_ 30-49% predicted; GOLD 4: FEV_1_/FVC < 0.70 and post-bronchodilator FEV_1_ < 30% predicted; MMRC: Modified medical research council dyspnea scoring system; PRISm: Preserved Ratio Impaired Spirometry (defined as FEV_1_/FVC ≥ 0.70 but with FEV_1_ < 80% predicted); SGRQ: St. George’s respiratory questionnaire score.  Adjusted Perc15 density: Cut off value in Hounsfield units (HU) below which 15% of all voxels are distributed on a lung CT scan (per convention, adjusted Perc15 density values are reported as the HU + 1000); Gas trapping (%): Percentage of lung voxels with a density less than -856 HU at end exhalation; % LAA-950: Percentage of CT low attenuation less than -950 HU at end-inspiration using Thirona software; Pi10: Square root of the wall area of a hypothetical airway of a 10-mm internal perimeter; % Segmental airway wall thickness: Percentage of the wall relative to the total bronchial area for the segmental airways; Upper/lower emphysema ratio: Log of the lung upper third to lower third ratio of emphysema.  P-values correspond to global P-values for comparisons across all 4 k-means clusters. P-values < 0.05 are italicized. | | | | | |

**Table S3.** Pairwise P-values between k-means clusters for annualized changes in FEV_1_ and adjusted Perc15 density

|  |  | RRS | ULE | AD |
| --- | --- | --- | --- | --- |
| Absolute annualized FEV_1_ changes | ULE | 0.6 | - | - |
|  | AD | *< 0.0001* | *0.004* | - |
|  | SE | 0.4 | 1.0 | *0.01* |
| Relative annualized FEV_1_ changes (percent changes from baseline) | ULE | *0.02* | - | - |
|  | AD | 1.0 | 0.04 | - |
|  | SE | *< 0.0001* | *< 0.0001* | *< 0.0001* |
| Absolute annualized adjusted Perc15 density changes | ULE | *< 0.0001* | - | - |
|  | AD | *< 0.0001* | 0.06 | - |
|  | SE | *< 0.0001* | 1.0 | 0.1 |
| Relative annualized adjusted Perc15 density changes (percent changes from baseline) | ULE | *< 0.0001* | - | - |
|  | AD | *< 0.0001* | *0.03* | - |
|  | SE | *< 0.0001* | *0.02* | *< 0.0001* |
| Absolute annualized changes were computed by subtracting Visit 1 values from Visit 2 values and dividing by the time in years between both visits for each subject. Relative annualized changes were calculated by dividing absolute annualized changes by Visit 1 values and multiplying by 100.  Per convention, adjusted Perc15 density values are reported as the HU + 1000.  Pairwise P-values were obtained using the Nemenyi test. P-values < 0.05 are italicized.  Abbreviations: RRS = Relatively resistant smokers; ULE = Upper lobe predominant emphysema; AD = Airway-predominant disease; SE = Severe emphysema. | | | | |

**Table S4.** Associations between clusters for absolute and relative annualized emphysema changes after adding scanner types as a covariate in the subgroup analysis with only subjects who underwent scans with the same scanner type between visits

|  | **K-means cluster** | **Univariable models** | | **Multivariable models** | |
| --- | --- | --- | --- | --- | --- |
|  |  | **Beta (Std Err)** | **P-value** | **Beta (Std Err)** | **P-value** |
| **Absolute annualized emphysema (adjusted Perc15 density) changes** | ULE | -0.66 (0.11) | *< 0.0001* | -0.58 (0.11) | *< 0.0001* |
|  | AD | -0.31 (0.09) | *0.0008* | -0.29 (0.09) | *0.003* |
|  | SE | -1.06 (0.11) | *< 0.0001* | -1.02 (0.12) | *< 0.0001* |
| **Relative annualized emphysema (% adjusted Perc15 density) changes** | ULE | -0.75 (0.12) | *< 0.0001* | -0.64 (0.13) | *< 0.0001* |
|  | AD | -0.31 (0.11) | *0.004* | -0.33 (0.11) | *0.003* |
|  | SE | -2.39 (0.13) | *< 0.0001* | -2.29 (0.13) | *< 0.0001* |
| Absolute annualized changes were computed by subtracting Visit 1 values from Visit 2 values and dividing by the time in years between both visits for each subject. Relative annualized changes were calculated by dividing absolute annualized changes by Visit 1 values and multiplying by 100. Negative values indicate worsening of the disease between visits.  Per convention, adjusted Perc15 density values are reported as the HU + 1000.  A total of 2,557 (from the 4,387 subjects with available Visit 1 and 2 adjusted Perc15 density values) had CT chest with identical scanner types in both Visits 1 and 2. Univariable linear regression models included only visit 1 k-means cluster assignment. Multivariable models also included adjustments for age, CT scanner type, sex, race, BMI, and smoking pack-years. The reference group was the relatively resistant smokers cluster (RRS) cluster.  Pairwise comparisons showed that the SE cluster had significantly greater absolute and relative emphysema changes than the RRS, ULE and AD clusters *(P-values < 0.05*), the ULE cluster had greater absolute and relative emphysema changes than the RRS and AD clusters *(P-values < 0.05)*, and the AD cluster had significantly greater absolute and relative emphysema changes than the RRS cluster *(P-values < 0.05)*.  P-values < 0.05 are italicized.  Abbreviations: Std Err = standard error; RRS = Relatively resistant smokers; ULE = Upper lobe predominant emphysema; AD = Airway-predominant disease; SE = Severe emphysema. | | | | | |

**Table S5.** Risks of COPD-related events and incident comorbidities between clusters

|  | **K-means cluster** | **Univariable models** | | **Multivariable models** | |
| --- | --- | --- | --- | --- | --- |
|  |  | **Hazard ratio (Std Err)** | **P-value** | **Hazard ratio (Std Err)** | **P-value** |
| **COPD exacerbations** | ULE | 1.48 (0.06) | *< 0.0001* | 1.35 (0.06) | *< 0.0001* |
|  | AD | 1.62 (0.05) | *< 0.0001* | 1.34 (0.05) | *< 0.0001* |
|  | SE | 3.96 (0.04) | *< 0.0001* | 2.98 (0.05) | *< 0.0001* |
| **CVD** | ULE | 1.38 (0.11) | *0.003* | 1.30 (0.11) | *0.02* |
|  | AD | 1.18 (0.09) | 0.08 | 1.12 (0.10) | 0.2 |
|  | SE | 1.55 (0.10) | *< 0.0001* | 1.25 (0.10) | *0.03* |
| **Diabetes** | ULE | 1.05 (0.12) | 0.7 | 0.99 (0.12) | 0.9 |
|  | AD | 1.97 (0.09) | *< 0.0001* | 1.44 (0.09) | *< 0.0001* |
|  | SE | 1.25 (0.11) | *0.04* | 1.31 (0.11) | *0.02* |
| Univariable Cox-proportional hazard models included only visit 1 k-means cluster assignment. The multivariable models for COPD exacerbations were also adjusted for COPD exacerbation history, age, sex, race, BMI, and smoking pack-years. The multivariable models for cardiovascular disease were also adjusted for age, sex, race, BMI, metabolic syndrome (defined as having at least 3 of the following conditions: BMI > 30, diabetes, hypertension, and high cholesterol) and smoking pack-years. The multivariable models for diabetes, all-cause mortality, cause-specific mortality, and mortality due to other causes were also adjusted for age, sex, race, BMI, and smoking pack-years. The reference group was the relatively resistant smokers (RRS) cluster.  Log-rank tests were used to compare clusters and multiple comparisons were corrected with the Benjamini-Hochberg method. The log-rank tests revealed that the SE cluster had higher risks for COPD exacerbations than the RRS, ULE, and AD clusters *(P-values < 0.05)*, and higher risks for CVD events than the RRS and AD clusters *(P-values < 0.05)*. The ULE cluster had higher risks for COPD exacerbations and CVD events than the RRS cluster *(P-values < 0.05)*. The AD cluster had higher risks for diabetes than the RRS, ULE, and SE clusters (*P-values <* *0.0001*), and higher risks of COPD exacerbations and CVD than the RRS cluster (*P-values <* *0.05*).  P-values < 0.05 are italicized.  Abbreviations: Std Err = standard error; ULE = Upper lobe predominant emphysema; AD = Airway-predominant disease; SE = Severe emphysema; CVD = Cardiovascular disease (defined as a composite endpoint of stroke, heart attack, coronary artery disease diagnosis, coronary artery bypass graft surgery, peripheral artery disease diagnosis, and/or cardiac angina). | | | | | |

**Table S6.** Risks of mortality between clusters

|  | **K-means cluster** | **Univariable model** | | **Multivariable model 1** | | **Multivariable model 2** | |
| --- | --- | --- | --- | --- | --- | --- | --- |
|  |  | **HR (Std Err)** | **P-value** | **HR (Std Err)** | **P-value** | **HR (Std Err)** | **P-value** |
| **All-cause mortality** | ULE | 1.91 (0.09) | *< 0.0001* | 1.73 (0.09) | *< 0.0001* | 1.48 (0.09) | *< 0.0001* |
|  | AD | 1.84 (0.08) | *< 0.0001* | 2.02 (0.08) | *< 0.0001* | 1.30 (0.08) | *< 0.0001* |
|  | SE | 5.87 (0.07) | *< 0.0001* | 4.34 (0.07) | *< 0.0001* | 1.64 (0.09) | *< 0.0001* |
| **COPD respiratory mortality** | ULE | 2.40 (0.41) | *0.03* | 2.12 (0.41) | 0.07 | 1.59 (0.41) | 0.3 |
|  | AD | 3.08 (0.34) | *0.001* | 3.77 (0.34) | *0.0001* | 1.51 (0.35) | 0.2 |
|  | SE | 50.1 (0.28) | *< 0.0001* | 34.7 (0.29) | *< 0.0001* | 5.75 (0.32) | *< 0.0001* |
| **CVD mortality** | ULE | 2.29 (0.26) | *0.002* | 2.23 (0.27) | *0.003* | 1.95 (0.27) | *0.01* |
|  | AD | 2.49 (0.23) | *< 0.0001* | 2.54 (0.23) | *< 0.0001* | 1.98 (0.24) | *0.004* |
|  | SE | 3.15 (0.23) | *< 0.0001* | 2.30 (0.24) | *0.0006* | 0.98 (0.30) | 1.0 |
| **Cancer mortality** | ULE | 2.00 (0.20) | *< 0.0001* | 1.81 (0.20) | *0.003* | 1.62 (0.20) | *0.02* |
|  | AD | 1.18 (0.19) | 0.4 | 1.28 (0.20) | 0.2 | 0.96 (0.20) | 0.9 |
|  | SE | 3.28 (0.17) | *< 0.0001* | 1.99 (0.17) | *< 0.0001* | 0.95 (0.22) | 0.8 |
| **Other mortality** | ULE | 1.27 (0.21) | 0.3 | 1.20 (0.22) | 0.4 | 1.09 (0.22) | 0.7 |
|  | AD | 1.37 (0.17) | 0.07 | 1.41 (0.18) | 0.06 | 1.03 (0.18) | 0.9 |
|  | SE | 2.3 (0.17) | *< 0.0001* | 2.06 (0.18) | *< 0.0001* | 0.95 (0.23) | 0.8 |
| Univariable Cox-proportional hazard models included only visit 1 k-means cluster assignment. Multivariable model 1 also included adjustment for age, sex, race, BMI and smoking pack-years. In multivariable model 2, the body mass index, airflow obstruction, dyspnea, and exercise capacity (BODE) index was also added as a covariate. The reference group was the relatively resistant smokers (RRS) cluster.  Log-rank tests were used to compare clusters and multiple comparisons were corrected with the Benjamini-Hochberg method.  The log-rank tests revealed that the SE cluster had higher risks of all-cause, COPD respiratory, cancer and other-causes related mortalities than the RRS, ULE, and AD clusters *(P-values < 0.05)*. The ULE cluster had higher risks of all-cause, COPD respiratory, CVD, and cancer mortalities than the RRS cluster *(P-values < 0.05)*. The AD cluster had higher risks of all-cause, COPD respiratory and CVD mortalities than the RRS cluster (*P-values < 0.05)*. The AD cluster had higher risks of all-cause, COPD respiratory and CVD mortalities than the RRS cluster, and higher cancer mortality than the ULE cluster *(P-values < 0.05)*.  Abbreviations: Std Err = standard error. ULE = Upper lobe predominant emphysema; AD = Airway-predominant disease; SE = Severe emphysema; CVD = Cardiovascular disease; HR = Hazard ratio; SE = Standard error.  P-values < 0.05 are italicized. | | | | | | | |

**The following 3 tables below are enclosed in separate Excel documents**

**Table S7.** SOMAscan plasma proteins associated to k-means cluster membership (univariable linear regression models on the left and multivariable linear regression models on the right). Covariates: k-means cluster assignment, age, sex, race, and current smoking status. In this file, there are three sheets for ULE vs. RRS, AD vs. RRS, and SE vs RRS comparisons and one sheet where we are listing the significant proteins (FDR 10%). The cluster following the “vs.” is the reference group. Abbreviations: RRS = Relatively resistant smokers; ULE = Upper lobe predominant emphysema; AD = Airway-predominant disease; SE = Severe emphysema.

**Table S8.** Differential gene expression between k-means clusters (covariates: k-means cluster assignment, age, race, sex, and current smoking status, white blood cell count proportions, and library prep batch). We corrected for multiple comparisons with the Benjamini-Hochberg method. In this file, there are 6 sheets for each pairwise comparison between the four k-means clusters. The cluster following the “vs.” is the reference group. Abbreviations: RRS = Relatively resistant smokers; ULE = Upper lobe predominant emphysema; AD = Airway predominant disease; SE = Severe emphysema.

**Table S9.** Significantly enriched gene ontology (GO) terms between k-means clusters (*weighted Fisher P-values < 0.005* and number of significant genes ≥ 3). In this file, there are 6 sheets for each pairwise comparisons between the four k-means clusters. The cluster following the “vs.” is the reference group. Abbreviations: RRS = Relatively resistant smokers; ULE = Upper lobe predominant emphysema; AD = Airway predominant disease; SE = Severe emphysema.
