## Supplementary figures and images for "Clustering-based COPD Subtypes Have Distinct Longitudinal Outcomes and Multi-omics Biomarkers"

### Figure S1

**Figure S1**

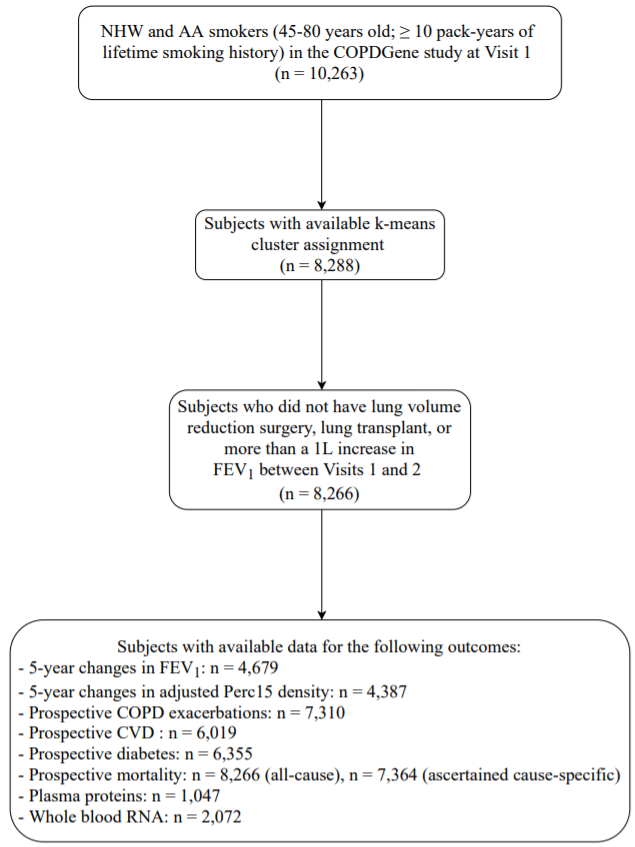

### Figure S2

**Figure S2**

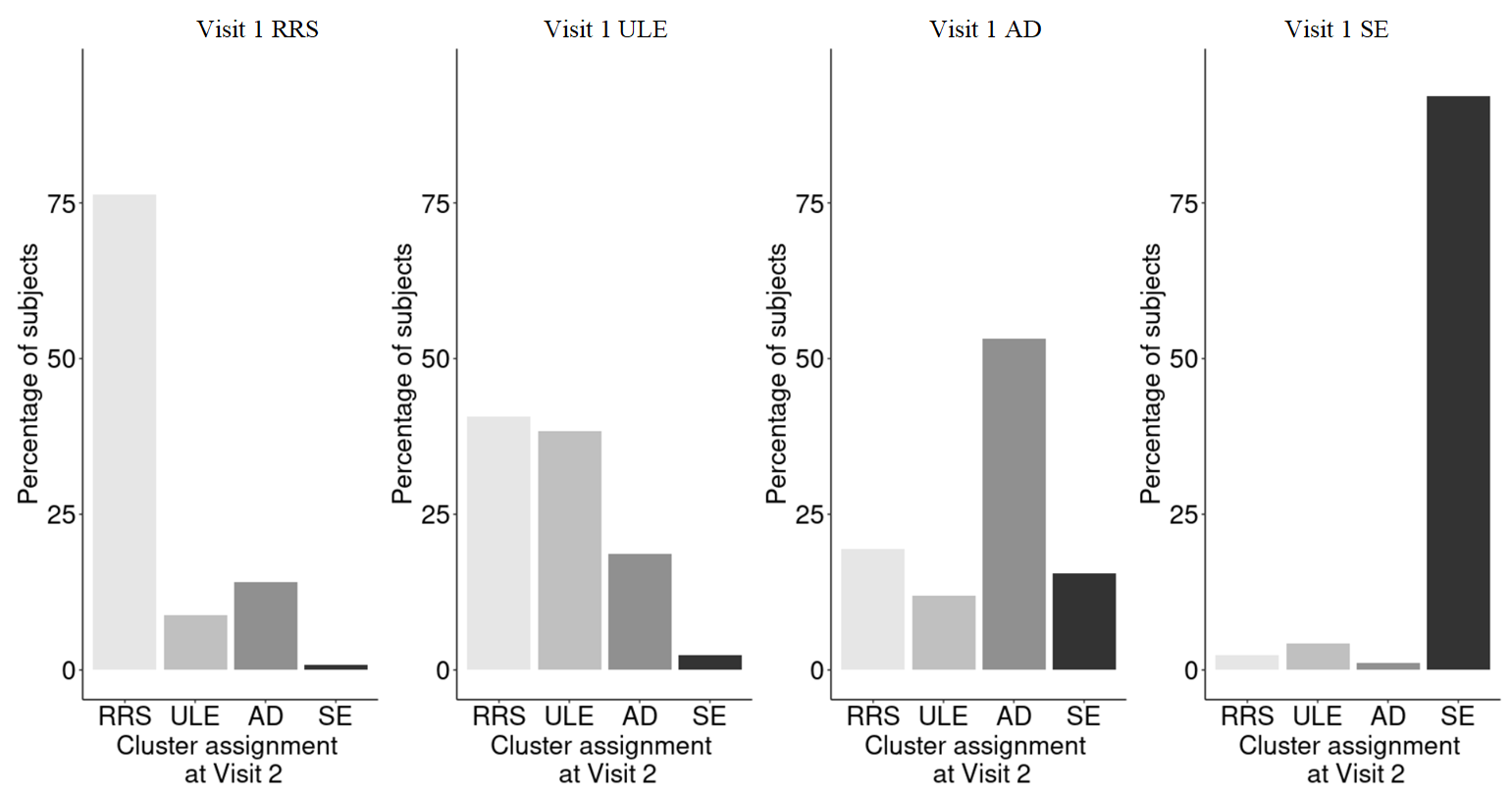

### Figure S3

**Figure S3**

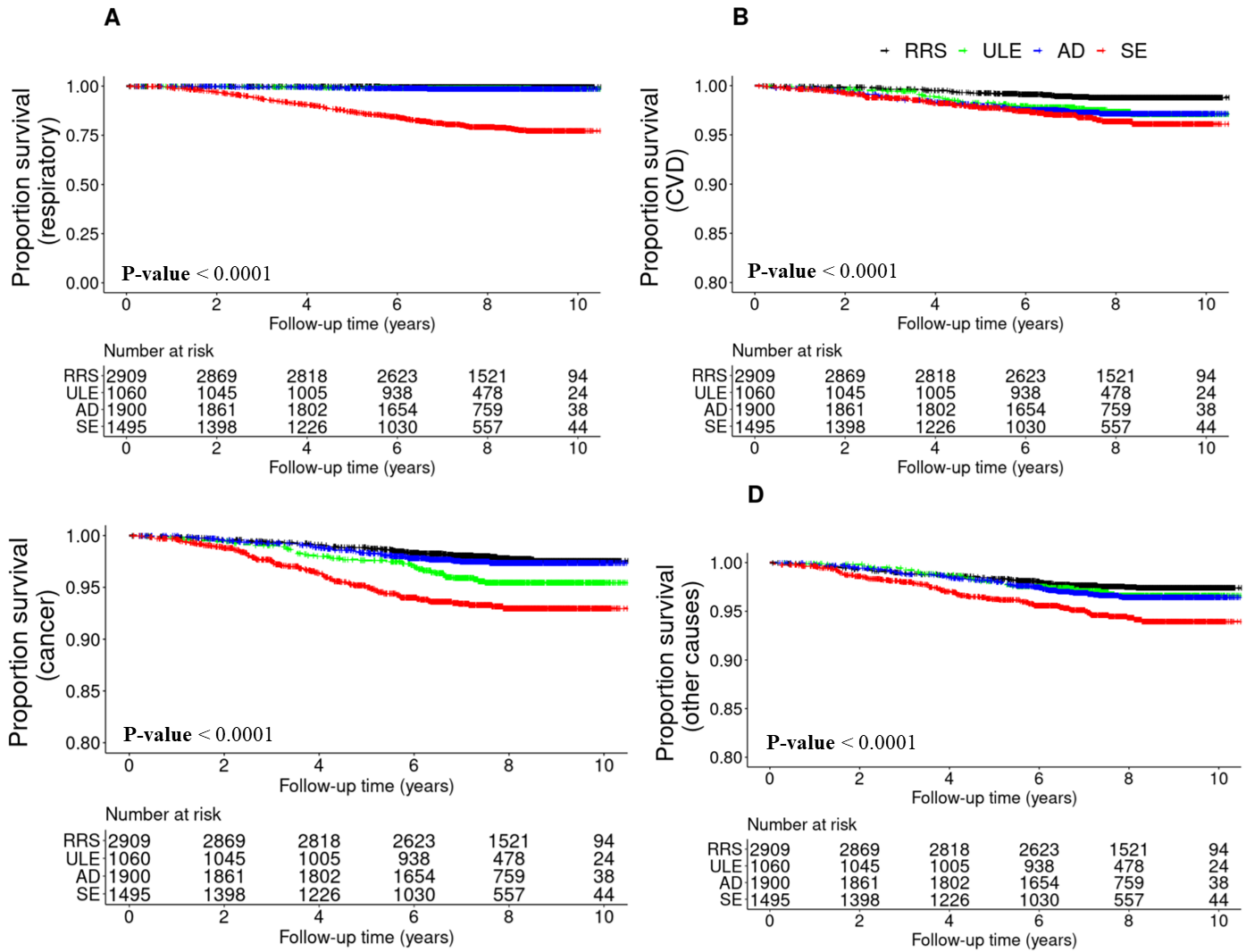

### Figure S4

**Figure S4**

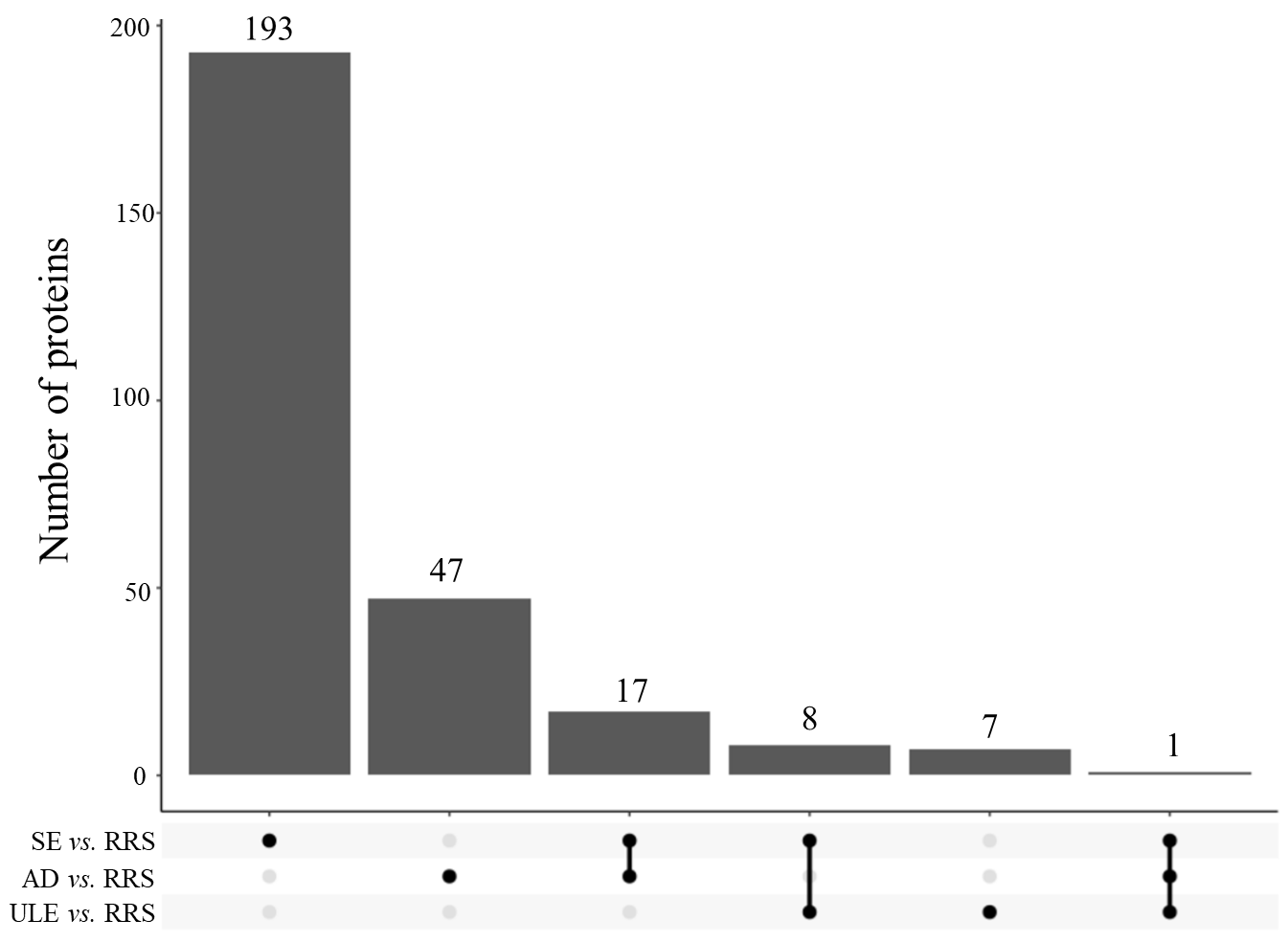
